# Mapping the genetic landscape of eight common cardiovascular diseases

**DOI:** 10.64898/2026.04.26.26351760

**Authors:** Cato Romero, Douglas Wightman, Sean J. Jurgens, Eva van Walree, Marre E. Corver, Poeya Haydarlou, Marijn Schipper, Connie R. Bezzina, Danielle Posthuma, Sophie van der Sluis

## Abstract

Cardiovascular diseases (CVDs) frequently co-occur, yet the shared genetic basis of cardiovascular multimorbidity remains unclear. We analysed common- and rare-variant genetic overlap across eight major CVDs using genome-wide and exome-wide association data from ∼1.7 million individuals in European and East Asian biobanks. Fifteen CVD pairs showed significant genetic correlations, with shared common-variant covariance explaining a modest proportion of phenotypic comorbidity. Genomic structural equation modelling identified three latent genetic clusters, while pleiotropic loci and genes frequently spanned cluster boundaries. Prioritised genes converged on atherosclerosis-related processes, myocardial structural and electrical programmes, and vascular-wall biology. In conditional analyses, body composition and metabolic traits consistently attenuated shared genetic liability, whereas circulating biomarkers showed smaller effects. For adequately powered traits, common-variant architecture was broadly similar between European and East Asian ancestries. These results define a shared genetic framework for cardiovascular multimorbidity centred on systemic risk factors and vascular biology.

---

Cardiovascular diseases (CVDs) are among the leading causes of mortality worldwide, posing a significant burden on healthcare systems and patients. Although each CVD has its own distinct clinical course, these conditions frequently manifest together (1–4). Patients diagnosed with multiple CVDs face a substantially higher risk of complications and greater challenges in treatment (5,6). Comorbid CVD cases often exhibit more severe outcomes and accelerated disease progression compared to patients with any single CVD condition alone (7–9), highlighting the need to understand the biological basis of cardiovascular multimorbidity.

Recent genetic studies show substantial heritable overlap among different CVDs (10,11). However, the genetic architecture underlying cardiovascular multimorbidity—how multiple CVDs cluster genetically and which biological pathways drive this overlap—remains poorly characterised. Longitudinal studies further show that common metabolic markers can predict progression from an initial CVD event to additional cardiovascular conditions (12), suggesting that shared biological mechanisms may contribute to the accumulation of CVDs within individuals.

In this work, we leverage genetic data of ∼1.7 million people from five large European and East Asian biobanks to investigate genetic overlap between eight prevalent CVDs—abdominal aortic aneurysm (AAA), aortic valve stenosis (AS), atrial fibrillation (AF), other conduction disorders (CD), heart failure (HF), ischemic heart disease (IHD), venous thromboembolism (VTE), and varicose veins (VV). By analysing both common and rare variants, we aim to identify loci, genes, functional annotations, and risk factors illuminating the prevalent comorbidity among CVDs.

## Methods

### Participant cohorts and phenotype definition

UK Biobank (UKB (13)) served as the primary discovery cohort for European ancestry (EUR) genome-wide and exome-wide association analyses (GWAS and ExWAS). GWAS replication used publicly available summary statistics from FinnGen (14), while ExWAS replication used data from the All of Us Research Program (15,16). Cross-ancestry analyses used publicly available GWAS summary statistics from Taiwan Precision Medicine Initiative (17) and BioBank Japan (18). Replication rather than meta-analysis was used to reduce biobank-specific ascertainment bias (18–21; **Figure 1**).

**Figure 1.**
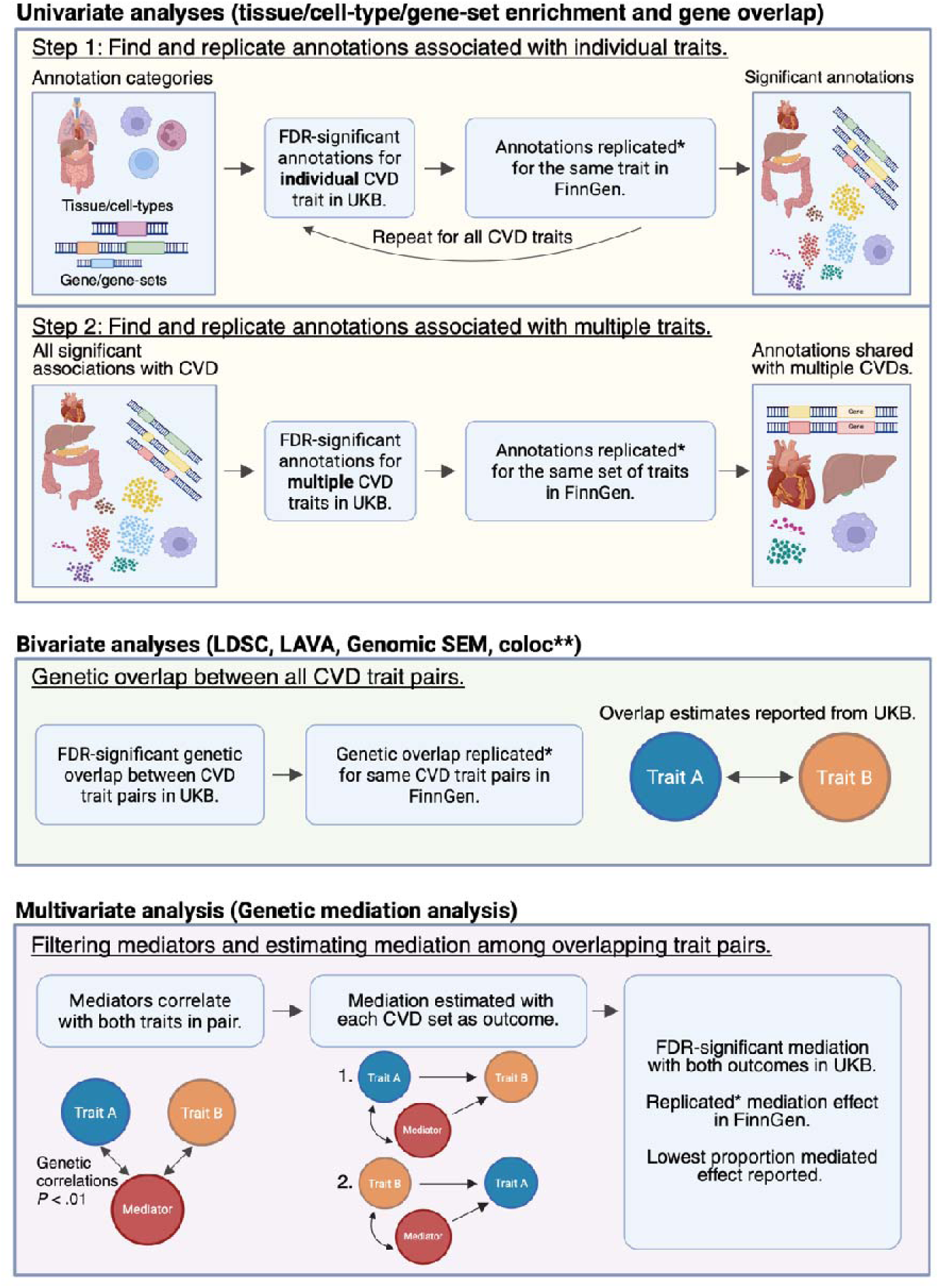
Overview of analytical workflow. The figure summarizes the univariate, bivariate, and multivariate analysis pipeline. Univariate analyses identify and replicate genes and functional annotations associated with individual CVD. Bivariate analyses test and replicate pairwise genetic overlap between traits. Multivariate analyses assess genetic mediation among trait pairs with replicated overlap. Discovery and replication cohorts are used throughout. When ‘replicated’ is marked with an asterisk, it refers to replication utilizing Bonferroni correction for multiple testing. Methods marked with two asterisks (**) follow the same conceptual pipeline but are based on Bayesian-methodology; thus, significance thresholds (e.g., p-values, FDR, Bonferroni correction) do not apply.

Access to the All of Us resource was granted by the All of Us Institutional Review Board, with analyses conducted under a data use agreement between Amsterdam UMC and the All of Us program. Use of UK Biobank resources was approved by the UK Biobank Research Ethics Committee, with UKB data accessed under approved applications 16406 and 176602.

Eight cardiovascular diseases (CVDs) were selected based on clinical relevance, effective sample size (N>10,000), and significant SNP-based heritability (*h^2^_SNP_*; α*_BON_* =.05/8). Cases in UKB were defined using ICD-9/ICD-10 codes across primary care, hospital, and death registry records (field 1712). Cohort-specific sample sizes, phenotype definitions, and additional details are provided in **Supplementary Table 1** and **Supplementary Note 1**. To prevent induced cross-trait similarity, individuals with more than one CVD diagnosis were reassigned such that each contributed to only one GWAS (**Supplementary Note 2** for a discussion; **Supplementary Figure 1–2**).

### Genome-wide association analysis and harmonization

GWAS for all eight CVDs were conducted in UKB participants of European ancestry using REGENIE v3.4.13 (23) with Firth correction, adjusting for age, genotyping array, and 20 ancestry principal components. Variants were filtered on minor allele frequency (≥0.01), missingness (≤0.05), and imputation quality (INFO ≥0.9), yielding approximately 8.5 million SNPs. Related individuals with discordant CVD diagnoses were excluded (**Supplementary Note 3**). Summary statistics were harmonized to hg19 using dbSNP (24) and restricted to SNPs present across all cohorts, yielding 4,637,379 shared variants across 22 autosomes and the X chromosome.

### Global genetic correlation and contribution to phenotypic comorbidity

*h*^2^_SNP_ and bivariate genetic correlations (*r*_g_) were estimated using linkage disequilibrium score regression (LDSC-v.1.0.1 (25)), with the 1000 Genomes EUR LD reference panel, restricted to HapMap3 SNPs. Discovery *r*_g_s were obtained in UKB and replicated in FinnGen using Bonferroni correction for FDR-significant pairs. For replicated pairwise *r*_g_s, the proportion of phenotypic comorbidity explained by genetic covariance was estimated as the ratio of LDSC-derived genetic covariance to tetrachoric phenotypic correlation in UKB, with standard errors derived using the delta method (25–26).

### Polygenicity and shared genetic architecture

Univariate and bivariate MiXeR (v1.3 (28)) analyses were applied to estimate trait-specific polygenicity and the proportion of shared trait-influencing variants between CVD pairs, reported as Dice coefficients. All analyses used the 1000 Genomes (v3) EUR reference panel to account for allele-frequency distributions and LD structure. The MHC region (hg19: chr6: 28,477,797–33,448,354 (29)) was excluded. Model fit was evaluated using Akaike information criterion (AIC), and poorly fitting models (negative AIC values), indicating that the data are not well captured by the assumed polygenic mixture model, were excluded from interpretation.

### Latent genetic factors structure

Genomic structural equation modelling (gSEM (30)) was applied to the LDSC genetic covariance matrix to identify latent genetic factors across the eight CVDs. Exploratory factor analysis tested one-to-four factor solutions; a three-factor confirmatory model was selected based on Akaike information criterion, comparative fit index, and standardized root mean square residual, and replicated using FinnGen GWAS summary statistics.

### Local genetic correlation analysis

Local genetic correlations were estimated using LAVA (v0.1.0 (31)) across 2,567 semi-independent ∼1 Mb LD blocks, including the X chromosome, using 1000 Genomes EUR as the reference panel. Regions with evidence of local SNP-heritability in more than one CVD (*P*<1×10DD) were taken forward to bivariate testing. LAVA adjusts for sample overlap using bivariate LDSC intercepts. Significant local correlations after FDR correction in UKB were replicated in FinnGen under Bonferroni correction. All tests were two-sided.

### Colocalisation of shared loci

Colocalisation analyses were performed using coloc (v5.2.3 (32)) for genome-wide significant loci identified with FUMA (33) that replicated across cohorts. Coloc evaluates five hypotheses: no association (H0), association with trait 1 only (H1), association with trait 2 only (H2), two distinct causal variants (H3), or a shared causal variant (H4). Loci were considered pleiotropic when the posterior probability for either H3 or H4 exceeded 0.8 in both UKB and FinnGen analyses, as H3 can indicate gene-level pleiotropy when distinct variants converge on the same locus.

### Gene prioritisation

Effector genes within shared loci were prioritised using FLAMES (v1.1.0 (34)), which integrates fine-mapped credible sets, PoPS scores (a gene prioritisation score integrating co-expression and functional annotations), and functional annotations. Fine-mapping was performed using SuSiE (v0.12.41 (35)) within PolyFun (36) with in-sample LD references from UKB and FinnGen (**Data availability**). PoPS (v0.2 (37)) scores were derived incorporating co-expression features and gene-tissue association statistics from MAGMA. Genes with a FLAMES precision score ≥0.75 were considered high-confidence effector genes; genes exceeding this threshold for the same locus in more than one CVD were classified as shared.

### MAGMA gene annotation analyses

Gene-based statistics from MAGMA were used to perform gene-set (MSigDB v2023.1Hs), GTEx tissue (v8), and GTEx cell-type enrichment analyses (37–39). Parallel analyses were conducted using gene-burden statistics from the ExWAS to identify rare-variant biological annotations. For each analysis type, significant annotations were first identified in UKB (FDR <5%) and replicated in FinnGen (Bonferroni correction); replicated annotations were then tested for pleiotropy across CVDs in UKB (FDR <5%) and replicated again in FinnGen (Bonferroni correction; **Supplementary Note 4**).

### Genetic mediation analysis

To assess whether genetic correlations between CVDs were partly explained by shared liability with other traits, mediation analyses were conducted using gSEM. Candidate mediators included 288 traits spanning cardiometabolic risk factors, socioeconomic measures, biomarkers, and 249 circulating metabolites. For each CVD pair with significant *r*_g_, mediators genetically correlated with both CVDs (P<0.01) were tested by comparing the marginal *r*_g_ between the CVD pair with the conditional *r*_g_ obtained after adjusting for the mediator trait; a significant difference indicates that the mediator significantly contributes to the shared genetic covariance, with significance determined under Bonferroni correction (α_BON_ =.05/number of traits tested; see (39)). Models were evaluated in both orientations to avoid directional bias, and the minimum proportion mediated across orientations is reported as a conservative estimate.

### Phenotypic mediation

Counterfactual phenotypic mediation analyses were conducted in UKB using the regmedint R package, adjusting for age and, for blood biomarkers, statin use (atorvastatin, simvastatin, rosuvastatin; (40)). Each mediator was phenotypically tested only if it had shown significant genetic attenuation in the corresponding CVD pair. Bonferroni correction was applied across mediators tested per CVD pair.

### Cross-ancestry replication

GWAS summary statistics from BioBank Japan and Taiwan Precision Medicine Initiative were used for East Asian analyses. Traits available in both cohorts (AAA, AF, IHD, HF, VV) were meta-analysed using inverse-variance weighted fixed effects in METAL (41), while remaining traits were analysed in Taiwan Precision Medicine Initiative alone. Subsequent analyses were restricted to traits with significant SNP-based heritability in East Asian samples. Cross-ancestry similarity in genetic architecture was assessed using Popcorn (42). Genetic correlations and mediation analyses were repeated in East Asian datasets using LDSC and gSEM. Pleiotropic loci identified in EUR analyses were evaluated in East Asian GWAS using the lead SNP with the highest posterior inclusion probability, tested under Bonferroni correction.

### Exome-wide association analysis

Gene-based burden tests were conducted in UKB and All of Us using whole-genome sequencing data and REGENIE (v4.1 (43)), restricted to coding variants on canonical transcripts of protein-coding genes. Variant annotation used VEP with LOFTEE, dbNSFP (v4.2), PrimateAI-3D, popEVE, and AlphaMissense (44–48). Two variant categories were considered: high-confidence protein-truncating variants and predicted damaging missense variants, combined into multiple masks varying by predicted pathogenicity and rarity thresholds (MAF <0.01, <0.001, and <0.00001 based on population-specific allele frequencies from gnomAD v2). Burden masks were tested in UKB and in All of Us, adjusting at minimum for ancestral principal components, age, age², and genetically inferred sex. Mask-level p-values were combined per gene using the Cauchy combination test, yielding both an overall and a rare-variant-specific gene-level p-value (MAF <0.001). Rare-variant pleiotropic associations were identified using the same staged discovery–replication framework applied in GWAS analyses, with discovery in UKB (FDR <5%) and replication in All of Us (Bonferroni correction; **Supplementary Note 5**).

## Results

### Common and rare variant association analyses

We conducted separate GWAS and ExWAS on eight CVDs in the UKB (abdominal aortic aneurysm (AAA), aortic valve stenosis (AS), atrial fibrillation (AF), conduction disorders (CD), heart failure (HF), ischemic heart disease (IHD), venous thromboembolism (VTE), and varicose veins (VV).

For an overview of univariate results see **Supplementary Note 6**. We investigated shared genetic liability at multiple levels between the CVDs, using UKB as the discovery dataset and FinnGen (GWAS-only) and All of Us (ExWAS-only) as replication cohorts (**Figure 1**). For downstream analyses, unless otherwise specified, point estimates refer to UKB-based results and are replicated in FinnGen/All of Us (see Supplementary Tables for all cohort results).

### Quantifying shared genetic liability among eight cardiovascular diseases

All eight CVD GWAS showed significant common SNP-based heritability (*h*^2^_SNP_) ranging from 4.17% (CD) to 21.35% (AS) (median = 13.40%; **Figure 2A; Supplementary Table 2**). Global genetic correlations (*r*_g_) were significant for 15 out of 28 CVD pairs ranging from 0.10 (AF–VV) to 0.66 (IHD–HF; **Figure 2B; Supplementary Table 3; Supplementary Figure 3**), indicating that several CVDs share genetic effects with similar effect direction contributing towards their *h*^2^_SNP_.

**Figure 2.**
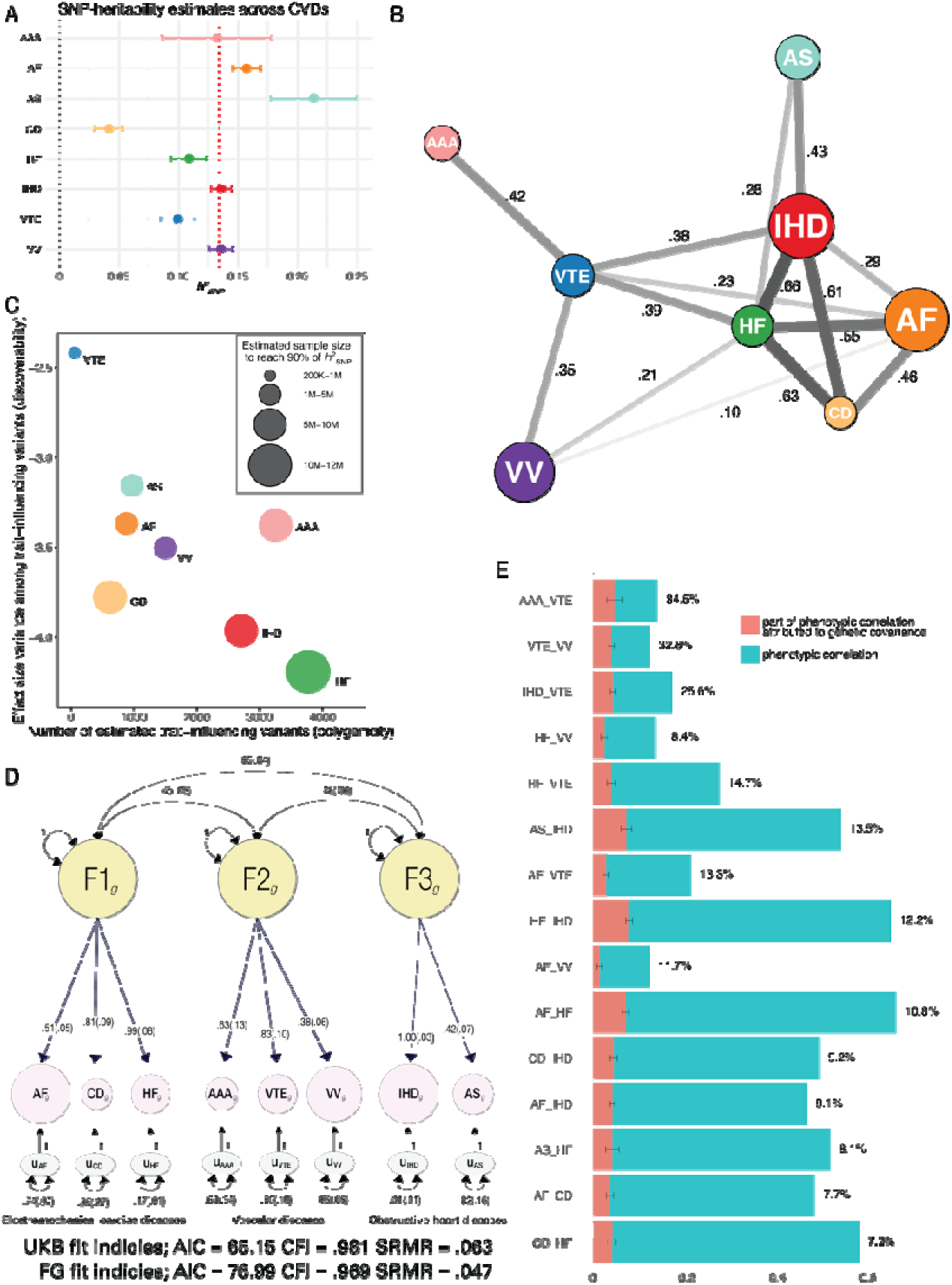
Genetic architecture and overlap across eight CVDs. (A) SNP-based heritability estimates for each CVD, with median indicated as the red dotted line. (B) Network of significant and replicated global genetic correlations; edge width reflects correlation strength and node size is weighted by SNP-heritability scaled by effective sample size (h^2^_SNP_ × sqrt(N_eff_)). (C) Univariate MiXeR results showing polygenicity, discoverability, and the estimated sample size required to capture 90% of SNP-heritability with genome-wide significant variants. (D) gSEM identifying three latent genetic clusters, with node sizes weighted by h^2^_SNP_ × sqrt(N_eff_). (E) Decomposition of phenotypic correlations, showing the total phenotypic correlation (blue-green) and the proportion attributable to common-variant genetic covariance (red).

To quantify shared genetic architecture between CVDs, we applied bivariate MiXeR. Across 11 CVD pairs with adequate model fit, MiXeR estimated that a substantial proportion of trait-influencing SNPs were shared (median Dice coefficient = 0.27; **Supplementary Figure 4**; **Supplementary Table 5**), despite relatively low polygenicity of CVD GWASs (median = 1,330 trait-influencing SNPs, compared with a median of 10,110 in other complex traits (49); **Figure 2C**; **Supplementary Table 4**).

gSEM was used to cluster CVDs based on their shared genetic covariance structure and revealed a well-fitting, well-interpretable genetic three-factor model (CFI = 0.98, SRMR = 0.06) grouping *electromechanical cardiac diseases* (AF, CD, HF), *vascular diseases* (AAA, VTE, VV), and *obstructive heart diseases* (AS, IHD; **Figure 2D; Supplementary Table 6**). We next examined how much of the phenotypic comorbidity between CVD pairs can be explained by their shared genetic covariance. We found that common variant-based genetic covariance explained a median of 12.8% (range = 7.2%–34.5%) of the phenotypic correlations (**Figure 2E; Supplementary Table 7**). Lastly, repeating analyses with sex-stratified GWAS analyses revealed no evidence for sex-specific differences in shared genetic liability or sex–SNP interactions across the eight CVDs, potentially due to low statistical power (**Supplementary Note 7**). Together, these findings indicate substantial shared genetic liability across CVDs, partly accounting for their phenotypic comorbidity and suggesting the involvement of genetic effects with broad cardiovascular relevance.

### Shared genomic regions and gene prioritization within pleiotropic CVD loci

Local genetic correlations (local *r*_g_s) across CVDs were estimated using LAVA within 2,567 predefined genomic regions. After filtering on univariate genetic signal, 507 regions showed evidence of association in more than one CVD. Subsequent 2,021 bivariate tests identified 30 significant local *r*_g_s across 13 CVD pairs, spanning 23 distinct genomic regions (**Figure 3; Supplementary Table 8**). Second, we applied colocalisation analysis on CVD GWS risk loci (from FUMA) using coloc (32). Out of 133 loci, coloc analyses identified 22 colocalized variants between CVD pairs spanning 9 loci; 5 of these loci fell within LAVA-identified regions (**Supplementary Table 9–10**). In total, LAVA and coloc identified 52 shared genetic signals between CVD pairs across 27 unique genomic regions. When summarized at the locus level, IHD emerged as the most connected phenotype, showing overlap with other CVDs in approximately half of all shared regions. At the pairwise level, overlap across multiple independent loci was most frequently observed for AAA–IHD (six regions) and AF–HF (four regions), while most other CVD pairs shared signals in one or two regions only.

**Figure 3.**
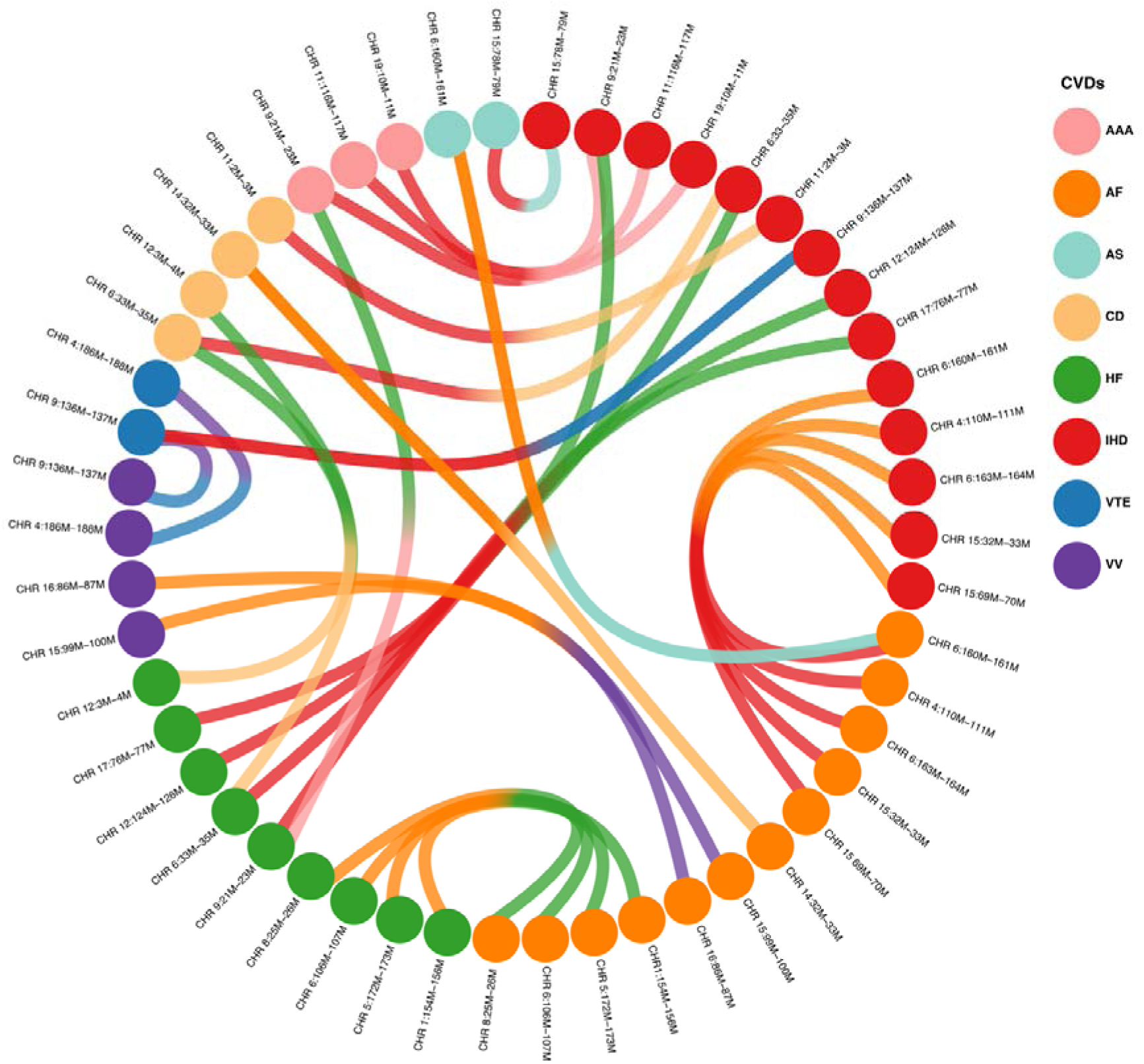
Shared genomic loci and shared gene predictions within pleiotropic loci across CVDs. Chord diagram of significant local genetic correlations from LAVA. Nodes are trait–locus instances, where the same genomic region (chromosome and base-pair interval) can appear once per trait; if a locus for one trait is shared with multiple other traits, multiple edges originate from that same node. Connected nodes represent the same locus shared between two traits with a significant local genetic correlation. Ribbons link trait pairs at each locus.

Having mapped shared genomic regions between CVD pairs, we used FLAMES to prioritize putative effector genes within these shared CVD genomic regions identified with LAVA and coloc. Of the 27 regions, 10 showed evidence for shared genes between CVD pairs (**Supplementary Table 11**). An additional three loci harboured likely shared effector genes among CVDs following inspection of locuszoom plots and previous functional validation published on corresponding CVDs in these regions (**Table 1**; for a discussion see **Supplementary Note 8**; locuszoom plots **Supplementary Figure 5–17**). Additionally, rare variant gene-based burden analyses identified five genes with associations across multiple CVDs: *PKD1* showed associations across AF, HF, IHD, and VTE; *TTN* was associated with AF, HF, and IHD (nominally to CD); *LMNA* was associated with AF, CD, and HF (nominally to IHD); *LDLR* was associated with AS, HF, and IHD; and finally, *SMAD6* showed associations with AS and CD (**Supplementary Table 12–16**). Of the five genes identified in rare variant burden analyses, two (*TTN* and *LDLR*) overlapped with genes prioritised from shared loci in the GWAS-based FLAMES analysis. Overall, mapping shared genomic regions and prioritising effector genes across CVDs identified several potential polygenic drivers underlying their shared genetic liability.

**Table 1.**
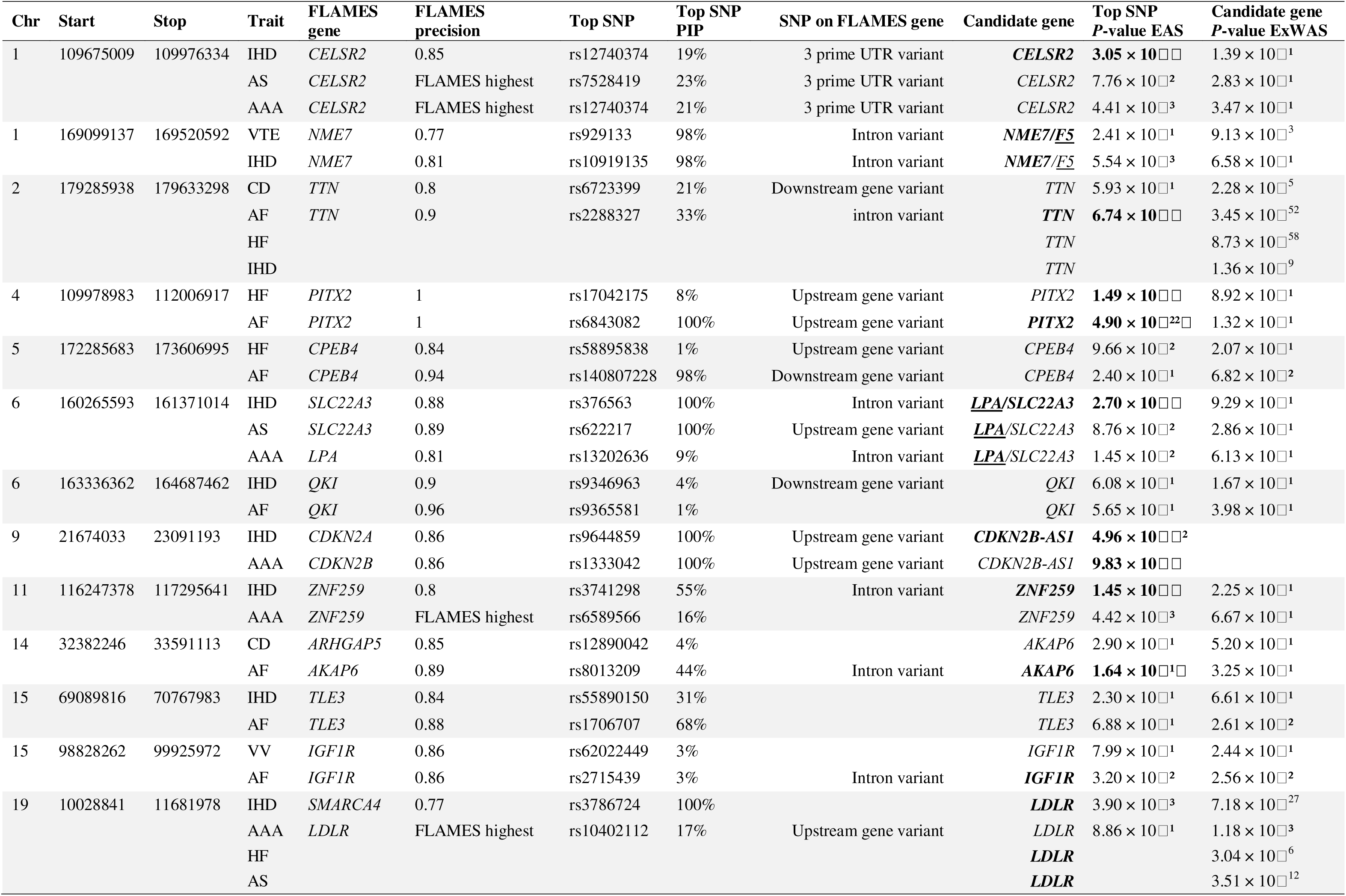
Overview of shared CVD loci and gene prioritisation. Note: FLAMES genes deviate from Candidate genes when additional evidence or limitations with FLAMES increase the likelihood that a different or multiple genes are potential effector genes. When Flames precision is below 0.75, the gene with the highest estimate was used and named ‘FLAMES highest’. SNP on FLAMES gene is obtained from Open Target Platform https://platform.opentargets.org. Candidate genes in bold are replicated in Qiao et al. 2025. Top SNP PIP indicates probability of SNP being the causal SNP in this locus. Top SNP P-value EAS is SNP based, not gene-based, while Candidate gene P-value ExWAS is gene-based, not SNP based. ‘Top SNP P-value in EAS’ values in bold are significant after bonerroni corrections (0.05/28).

### Shared biological annotations from CVD GWAS and ExWAS

We next sought to contextualize shared genetic vulnerability across CVDs by identifying biological annotations—tissues, cell types, and pathways—implicated across multiple CVD GWASs using MAGMA (**Supplementary Table 17**). Across 54 human tissues, we identified eight tissues associated to more than one CVD using gene-property analysis, including cardiovascular tissues (*coronary / tibial / aorta artery* and *atrial heart appendage*), gastrointestinal system (*sigmoid colon, oesophagus muscularis / gastroesophageal junction*), and *the uterus*. Most shared associations occurred between AF and VV, but overlap was also observed for IHD and HF (**Figure 4A; Supplementary Table 18**). Next, we evaluated 500 cell types from multiple human tissues (50), of which eight cell-types were associated with more than one CVDs. Among these, vascular-related cell-types—*endothelial cells* and *pericytes*—were implicated across multiple tissues (*pericytes in oesophagus, lung, and prostate; endothelial cells in liver heart* and *pituitary*). In addition, *heart mural cells* and liver-associated cell types (*fibroblasts* and *cholangiocytes*) showed significant associations with multiple CVDs (majority between VV, AF and IHD; **Figure 4B; Supplementary Table 19**).

**Figure 4.**
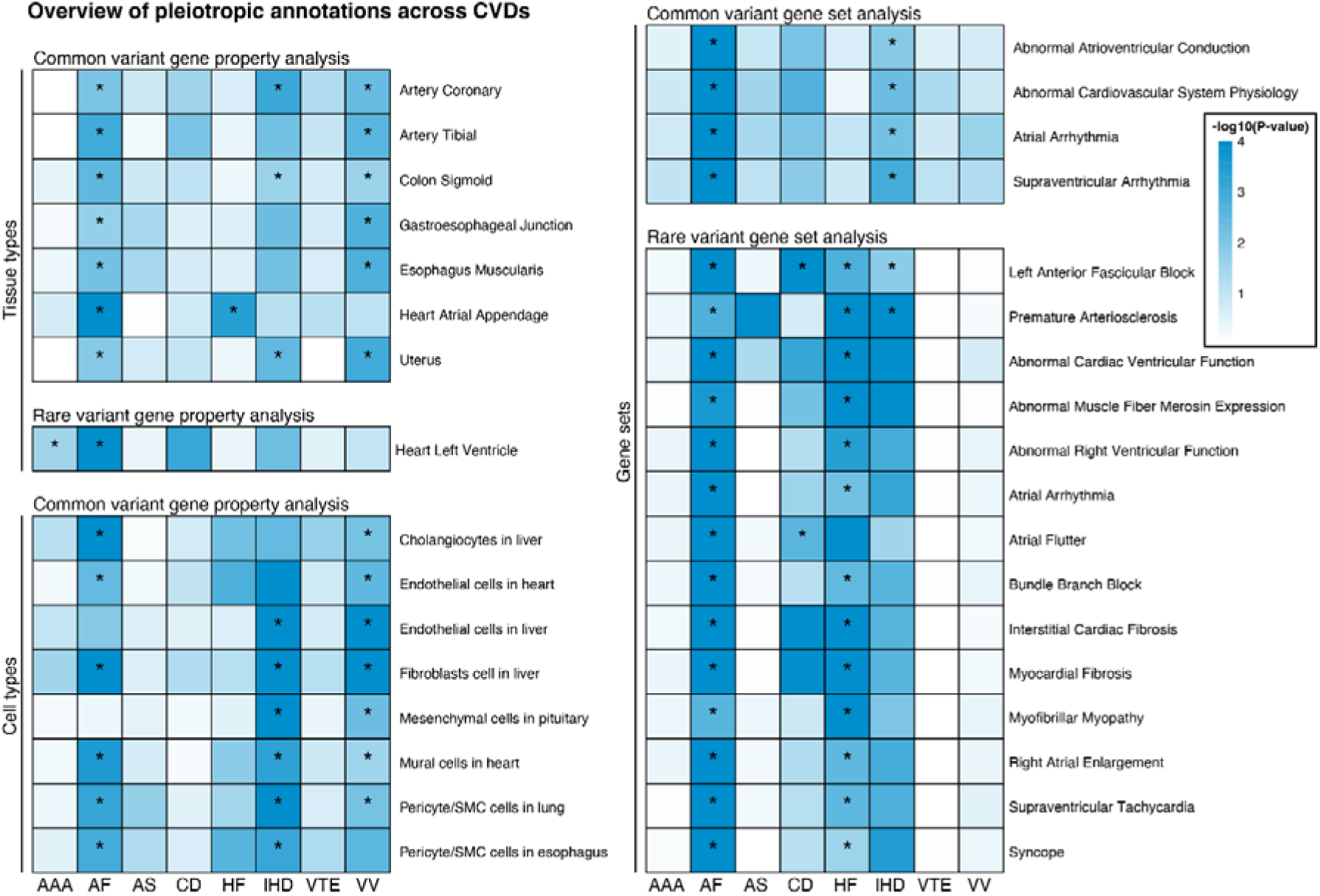
Pleiotropic gene-set, tissue, and cell-type enrichments across cardiovascular diseases. Heatmaps show MAGMA enrichment results for common-variant and rare-variant gene sets, as well as common-variant gene-property analyses for tissues and cell types across cardiovascular diseases. Color intensity reflects −log10(p-value), with darker blue indicating stronger evidence of enrichment. Asterisks denote annotations that replicate in FinnGen for the same trait–annotation pair. Only annotations showing overlap across at least one cardiovascular disease are displayed

We conducted gene-set analysis across 17,023 curated pathways and identified four gene-sets associated with more than one CVD; *supraventricular arrhythmia, atrial arrhythmia, abnormal cardiovascular system physiology,* and *abnormal atrioventricular conduction* (all between AF and IHD; **Figure 4C; Supplementary Table 20**). Parallel gene-property and gene-set analyses were performed using gene-burden associations from CVD ExWASs. No cell types were associated with more than one CVD; however, *heart left ventricle* expression was associated with both AAA and AF. In total, 14 gene sets were shared across CVDs in the ExWAS analyses. Notably, *left anterior fascicular block* was shared among AF, HF, IHD, and CD, and *premature arteriosclerosis* was shared among AF, HF, and IHD. The remaining 12 shared gene sets were predominantly observed between AF and HF (**Supplementary Table 21–23**). Together, these results suggest potential biological pathways and tissue/cell types through which CVD effector genes may influence shared cardiovascular pathology.

### Investigating CVD-shared risk factors using genetic mediation analysis

Having established widespread global genetic correlations between CVDs, we next examined whether this shared genetic liability is specific to CVD pairs or can be partly explained by genetic effects shared with a third trait. To this end, we applied genetic mediation analysis within a gSEM framework to assess whether the genetic liability of candidate risk factors attenuates pairwise CVD genetic correlations. Among 288 candidate risk factors (full list in **Supplementary Table 24**), 80 showed genetic correlations with both CVDs in at least one of the 15 significantly correlated CVD pairs and were carried forward for mediation analysis.

After correcting for multiple testing, genetic conditioning on body-mass index (BMI) or waist-hip ratio (WHR) resulted in the most widespread attenuation of CVD pair *r*_g_s, being significant in 10/15 and 6/15 CVD pairs, respectively. These effects were small to moderate across pairs (median reduction = 14%, range: 7% (CD–IHD), 26% (AF–VV); **Figure 5A**). Type 2 diabetes showed similar conditional effects in 5/15 CVD pairs (median = 14%, range: 11% for IHD–HF to 27% for AS–HF), followed by personal history of alcoholism being significant in 4/15 CVD pairs (median = 28%, range: 17% for IHD–VTE to 31% for AF–VTE). Hypertension was significant in three CVD pairs (18% for IHD–HF, 22% for IHD–CD, and 30% for IHD–AF), asthma in three CVD pairs (7% for AF–IHD, 11% for IHD–VTE, and 13% for AF–VTE), and smoking in one CVD pair (9% for AF–VTE). Conditioning on circulating blood metabolite markers yielded 30 significant attenuation effects across 6/15 CVD pairs (median = 9%, range: 6% for IHD–AF, 24% for AF–VV), with the majority (20/30) observed between IHD–HF (for full results see **Supplementary Table 25** and **Supplementary Figure 18–19**).

**Figure 5.**
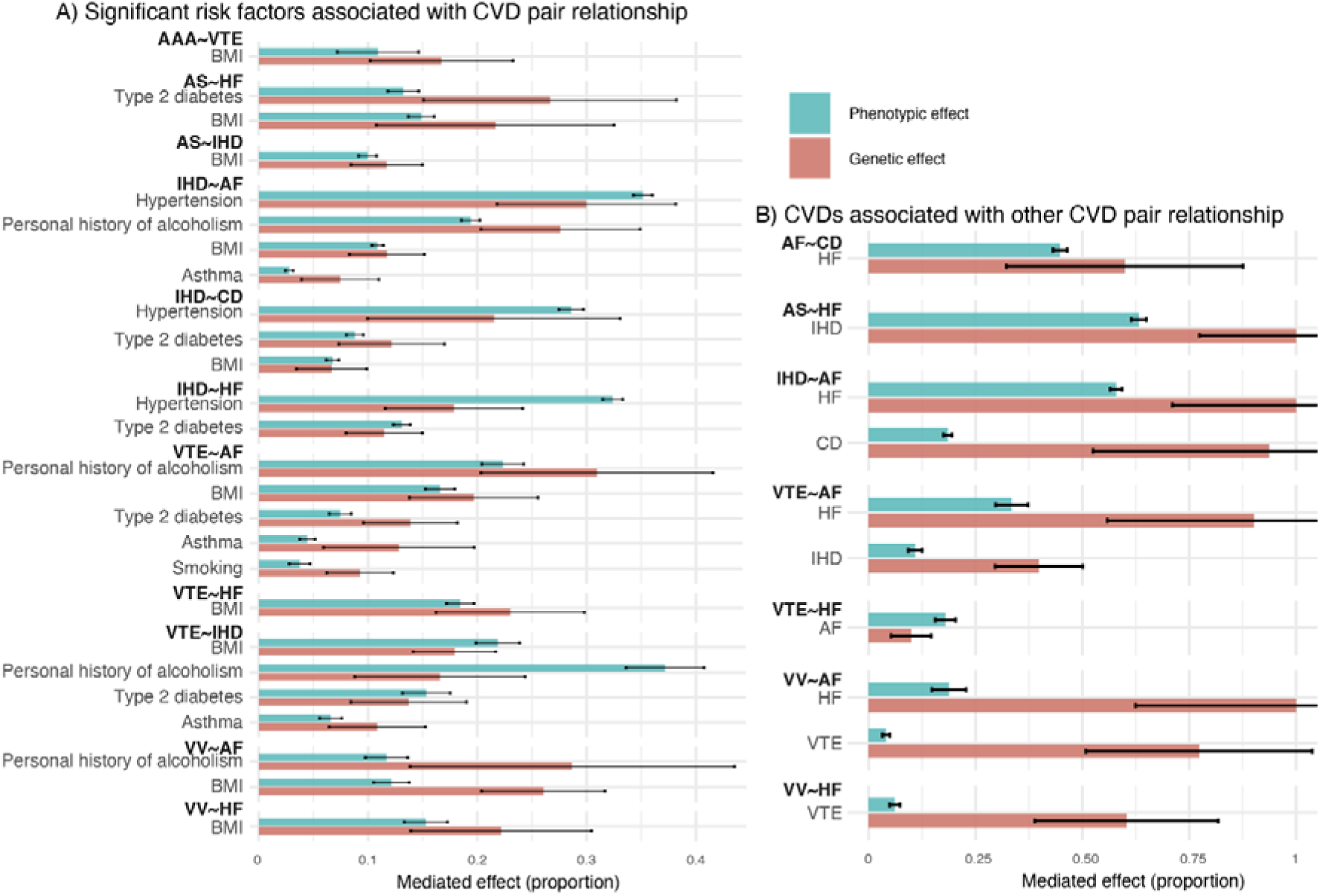
Genetic and phenotypic mediation of cardiovascular disease (CVD) pair relationships. (A) Significant mediator traits associated with specific CVD pair relationships. Bars show the proportion of the association mediated, estimated at the phenotypic level (blue) and genetic level (red). (B) Mediation effects of other CVDs on CVD pair relationships, shown analogously. For all mediators, mediation was estimated in both model orientations (A→B and B→A), and the minimum proportion mediated across orientations is shown as a conservative estimate. Error bars indicate standard errors

Conditioning on CVDs themselves as mediators between other CVD pairs revealed instances of near-complete attenuation, indicating that the observed genetic correlation was largely attributable to genetic effects shared with a third CVD rather than being unique to the original pair. Specifically, conditioning on HF between AF and IHD, VTE, or VV reduced the genetic correlation by more than 90%, while conditioning on IHD between AS–HF resulted in a complete attenuation of *r*_g_. Reductions exceeding 50% were also observed when conditioning VV–AF and VV–HF on VTE, AF–CD on HF, and AF–IHD on CD (**Figure 5B**).

At the phenotypic level, significant mediation effects were observed for all risk factor traits that had shown significant genetic attenuation, although effect sizes sometimes differed substantially (**Figure 5A–B**; **Supplementary Table 26**). We additionally performed genetic mediation analysis on the 30 significant local *r*_g_ using multivariate LAVA, revealing small to moderate attenuation effects for different serum fatty acid markers in 9/30 local *r*_g_s; however, none survived multiple testing correction (**Supplementary Note 9; Supplementary Figure 20; Supplementary Table 27**).

### Replication in East Asian genetic ancestry of available CVDs

To evaluate whether findings from EUR GWAS generalize to non-EUR populations, we repeated analyses using EAS-based GWAS of the same or closely related CVD traits from Biobank Japan and Taiwan Precision Medicine Initiative (case definitions and between-cohort *r*_g_s in **Supplementary Table 1**). Overlapping traits between EAS–cohorts were meta-analysed (AAA, AF, IHD, HF, and VV) using METAL. Among traits with significant SNP-heritability (AAA, AF, HF, and IHD), no significant deviation from unity was observed for either genetic impact correlation (similar effect sizes accounting for differences in MAF) or genetic effect correlation (similar effect sizes not accounting for differences in MAF) when compared with EUR-based GWAS using Popcorn (42), indicating no evidence for divergence in common-variant architecture across ancestries (**Supplementary Table 28**). Genetic correlations among these four EAS-based CVDs were significant except AAA–AF and AF–IHD (**Supplementary Table 29**). Genetic mediation analysis showed comparable attenuation effects to those observed in EUR ancestry: conditioning on smoking resulted in approximately 20% attenuation of genetic correlations for AAA–IHD and AAA–HF, while conditioning on type 2 diabetes resulted in approximately 10% attenuation for HF–AF and HF–IHD (**Supplementary Table 30**). Finally, of the 28 EUR-based pleiotropic CVD associations in **Table 1**, 15 showed nominal association in EAS-based GWAS of the corresponding traits, replicating pleiotropy for 5 of 13 loci. After correcting for multiple testing, 9 associations remained significant, corresponding to pleiotropic replication of 2 of 13 loci. Overall, several cross-CVD genetic signals identified in EUR ancestry were directionally consistent and suggestively replicated in EAS ancestry, although limited power constrained comprehensive replication across all traits and CVD pairs.

## Discussion

Cardiovascular diseases (CVD) frequently co-occur, yet the biological mechanisms underlying this comorbidity remain unresolved. In this study, we systematically mapped shared genetic effects contributing to comorbidity across a broad spectrum of CVDs by integrating common- and rare-variant association analyses with locus-level overlap, biological annotation, genetic mediation modelling, and cross-ancestry replication. Extending previous multivariate CVD GWAS-overlap studies, we examined a wider range of comorbid phenotypes, incorporated rare coding and X-chromosome variation, and contextualized shared genetic liability using mediation and cross-ancestry analyses. Together, these analyses provide a genetic framework for understanding why common CVDs cluster and which biological processes underpin this clustering (results overview in **Figure 6** and **Table 2**; pairwise discussion of CVD overlap in **Supplementary Note 10** and **Supplementary Table 31**).

**Figure 6.**
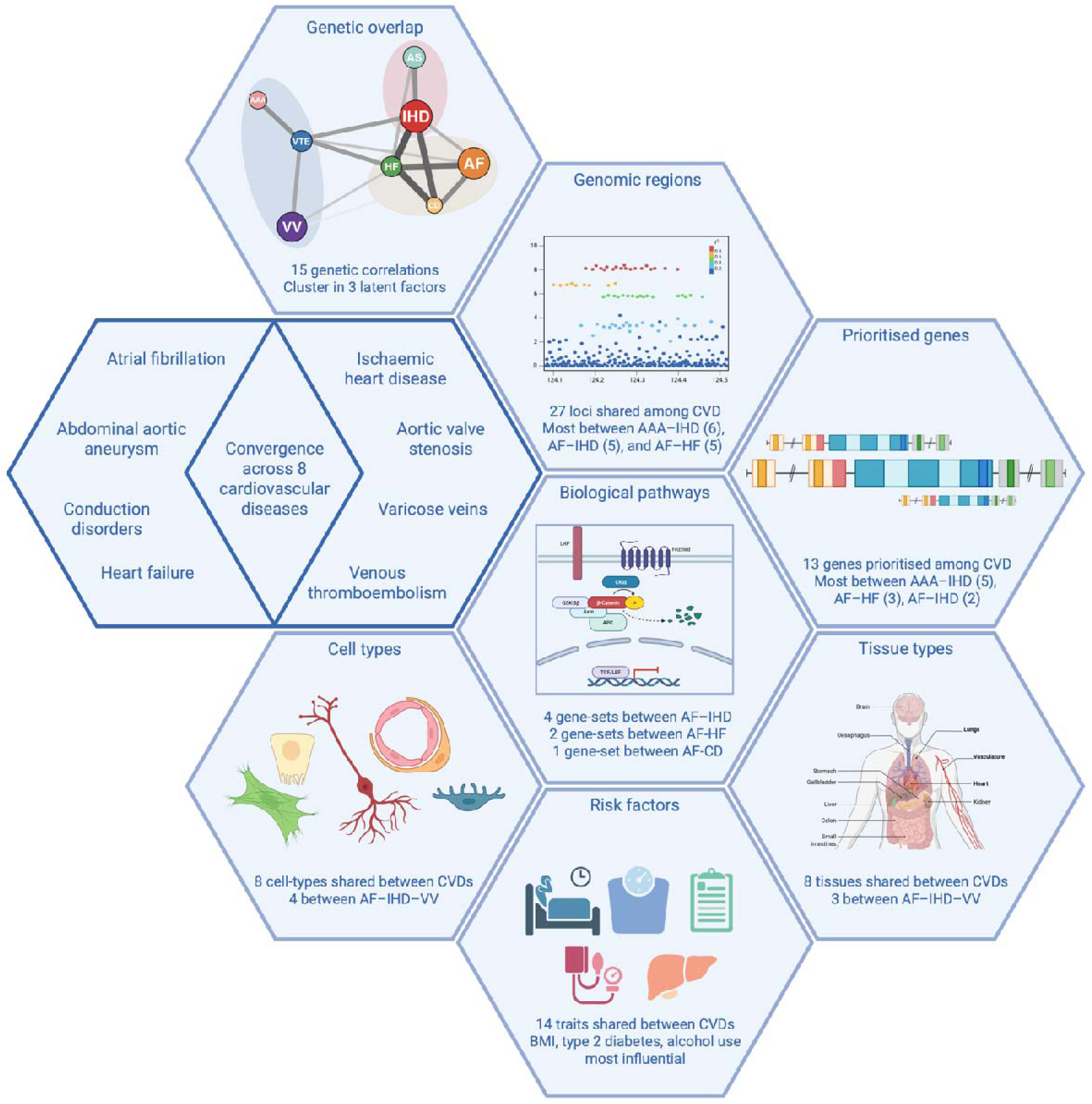
Schematic summary of all shared genetic overlap among eight CVDs. This figure summarises genetic associations across eight CVDs from genetic correlations, shared genomic regions, genes, pathways, tissue and cell enrichments and implicated risk factors identified in this study. Note: images are meant to be illustrative and not represent actual pathways identified in the analysis. Created with BioRender.com (see Acknowledgements).

**Table 2.**
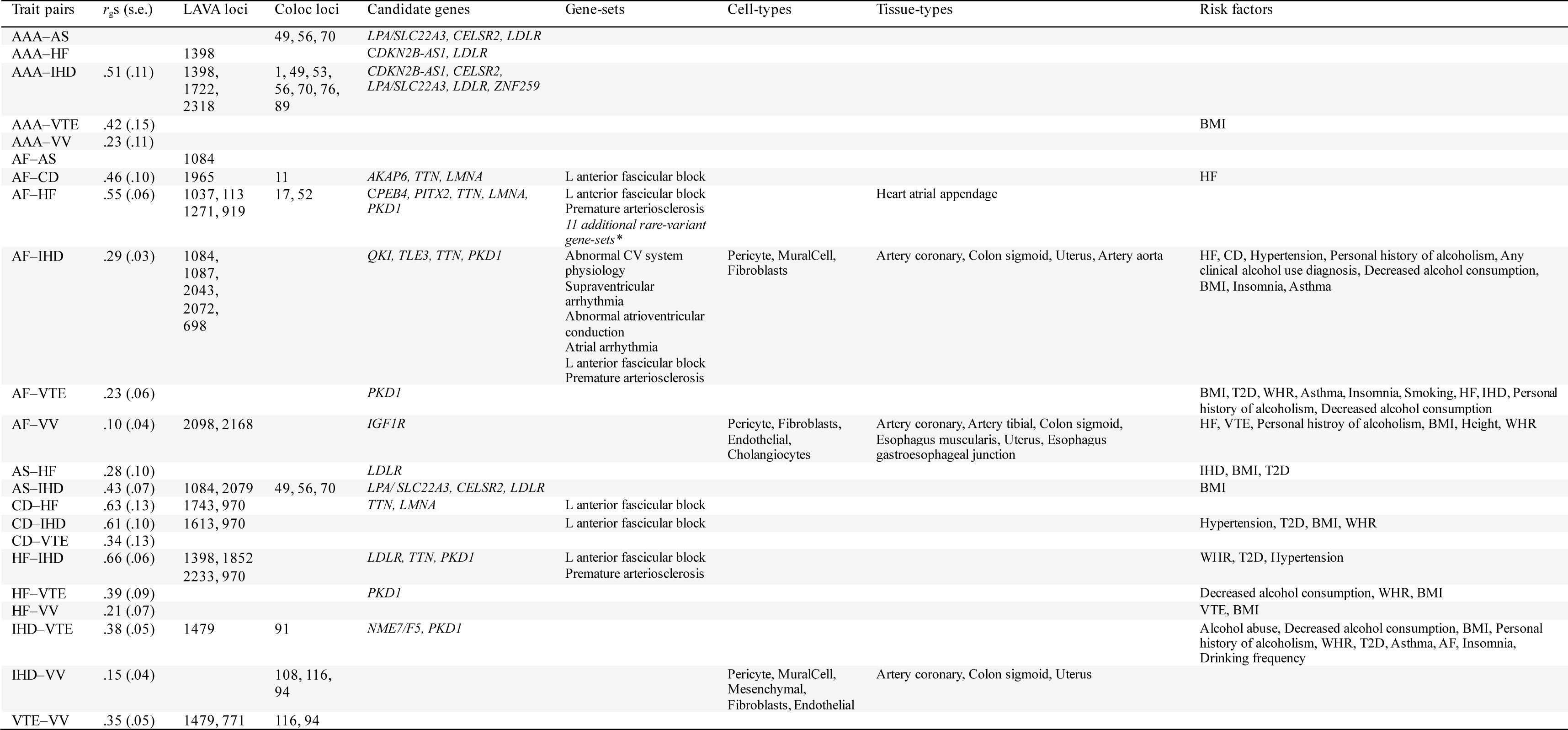
Genetic overlap across CVD pairs. Note: Shown are CVD pairs with evidence of shared genetic architecture across one or more analyses, including global genetic correlations, shared loci, candidate genes, gene-sets, cell or tissue enrichment, and genetic mediation by risk factors.

Latent factor modelling identified three clusters of CVDs based on their shared genome-wide genetic covariance, representing latent axes of genetic liability. The first cluster—comprising AF, CD and HF—captures electromechanical cardiac dysfunction, linking abnormalities in cardiac electrophysiology, conduction, and myocardial remodelling (51–53). The second cluster, including AAA, VTE, and VV, reflects vascular integrity and thrombosis spanning arterial (54) and venous beds (55–58). The third cluster groups IHD and AS, conditions characterised by progressive flow-limiting obstruction driven by atherosclerotic and calcific processes (59–62).

While this structure provides a useful conceptual organisation, several diseases—most notably HF—are biologically heterogeneous, such that a single-cluster assignment represents a necessary simplification. However, allowing traits to load on multiple factors did not improve model fit, likely because shared genetic influences across disease domains are already captured by the correlations between the three latent factors (0.45–0.65). The factor structure should therefore be interpreted as grouping CVDs with the most similar polygenic architecture, while acknowledging substantial genetic overlap across clusters.

Interestingly, shared genetic signals were predominantly observed between traits belonging to different latent factors. While the gSEM framework is based on *global* genetic covariance—an approach that is relatively robust to differences in GWAS power and captures broad patterns of shared liability—our locus-, gene-, and annotation-level analyses interrogate *local* genetic architecture and are therefore more sensitive to power and trait-specific genetic architecture. Across these local analyses, approximately 60% of shared loci and 70% of prioritised genes involved cross-factor trait pairs, and a similar pattern was observed for shared biological annotations. This organisation indicates that cross-CVD genetic architecture is structured around broad cardiovascular biological processes rather than factor-specific mechanisms. At the same time, detection of overlap was strongly influenced by statistical power: shared loci and annotations were most frequently observed among well-powered traits such as IHD, AF, and VV (**Figure 1B–D**), even when global genetic correlations between these traits were modest (63). This suggests that the apparent centrality of certain traits may partly reflect sample size and SNP-based heritability rather than biological exclusivity. Notably, gene overlap was substantially more pronounced for common-variant GWAS than for rare-variant ExWAS, consistent with recent work showing that common variants tend to exert more pleiotropic, system-wide effects, whereas rare coding variants are generally more trait-specific (64). This distinction has implications for therapeutic prioritisation, as rare-variant genes may reflect disease-specific mechanisms while common-variant genes often implicate cross-trait biology.

At the gene-level, several prioritised loci have well-established roles in cardiovascular biology. Across analyses, we identified 13 loci with evidence for shared genes between CVD pairs, 5 of which could not be unambiguously resolved by automated gene prioritisation due to non-coding effects, complex linkage disequilibrium, rare causal variants, or discordance between statistical prioritisation and biological plausibility (detailed discussion in **Supplementary Note 8**). We assessed replication against a recent cross-CVD study (10) that included AF, IHD, HF, and VTE. Across these four overlapping traits, our analyses yielded 21 gene–trait associations, of which 14 were reported in the previous study. Associations not observed in the previous study were confined to a small subset of cardiac-structural genes in HF (*TTN, PITX2, CPEB4*) and regulatory genes in AF and IHD (*QKI, TLE3*), potentially reflecting differences in study design and locus-discovery methodology. Notably, many previously reported genes also showed additional pleiotropic effects in CVDs examined only in the present study, including AS, AAA, CD, and VV, extending gene relevance across arterial, venous, and valvular disease.

Functionally, prioritised genes clustered into three partially overlapping categories. First, several loci implicated systemic lipid and circulating exposure pathways (*LDLR*(*65*)*, LPA*(*66,67*)*, CELSR2*(*68,69*)*, ZNF259*(*70*)*, SLC22A3*(*71*)). These genes were primarily detected in shared genetic signals involving IHD, AAA, and AS consistent with established roles of lipid metabolism and circulating risk factors in atherosclerotic and broader vascular disease. Second, genes related to cardiac structural, electrophysiological, and calcium-handling programs—including *TTN*(72,73)*, AKAP6*(74–76)*, PITX2*(77,78), *LMNA*(*79,80*)—highlighted shared genetic contributions among traits within the electromechanical cardiac cluster, specifically AF, CD, and HF. Third, a substantial fraction of prioritised genes were implicated in regulatory and signalling processes affecting vascular wall biology. These included RNA processing and cell-state control (*QKI*(81,82)*, CPEB4*(83–85)*, TLE3*(86)), cell-cycle and proliferative regulation (*CDKN2B-AS1*(87,88)), endothelial flow sensing and mechanotransduction (*NME7*(89,90), *PKD1*(*91,92*)), coagulation–endothelium interactions (*F5*(93,94)), and growth-factor and developmental signalling influencing vascular remodelling (*IGF1R*(95,96), *SMAD6*(97,98)). Notably, *SMAD6*, an inhibitory regulator of BMP signalling, emerged as a shared rare-variant burden gene between AS and CD, extending its established role in congenital cardiovascular defects to include conduction-related phenotypes (99). Except for *CDKN2B-AS1*, these genes were shared between AF and at least one additional CVD (VV, VTE, IHD, or HF), representing the gene group with the broadest cross-CVD involvement. Consistent with this concentration of shared genetic effects in vascular-wall regulatory pathways and their involvement across multiple CVDs, cell-type enrichment analyses revealed convergence on endothelial cells together with pericytes, smooth muscle cells, and stromal components across IHD, AF, and VV, supporting a common vascular-wall biological substrate underlying clinically distinct cardiovascular diseases.

Genetic mediation analyses further clarified the sources of shared liability. Body composition traits (BMI and WHR) showed the most consistent attenuation of genetic correlations across CVD pairs, in line with their established role as broad cardiovascular risk factors (100). Type 2 diabetes and alcohol-related traits exerted more selective but sometimes substantial effects, whereas hypertension—despite its central role in CVD risk—showed a genetic mediating role in only three CVD pairs, suggesting that its contribution to shared genetic liability across CVDs may be more phenotype-specific than that of adiposity-related traits, or may partly operate through non-genetic or downstream mechanisms. Circulating blood biomarkers generally exhibited modest attenuation effects, primarily between IHD and HF, suggesting that widespread metabolic perturbations rather than highly specific biomarkers contribute to shared genetic risk. As several mediating traits are modifiable, part of the comorbidity may operate through shared, modifiable cardiovascular risk factors, indicating potential for cross-disease prevention.

The strongest mediation effects were observed when conditioning CVDs on one another, with near-complete attenuation of genetic correlations in several trait pairs. For example, conditioning the genetic correlation between IHD and AF on HF resulted in complete attenuation, suggesting that genetic liability shared between IHD and AF also involves genetic effects that contribute to liability for HF. Because temporal ordering was not modelled, these findings do not imply causal directionality. Rather, they are compatible with models in which a substantial component of comorbidity between specific CVD pairs reflects vertical pleiotropy, whereby genetic liability to one cardiovascular condition increases susceptibility to multiple others, as opposed to horizontal pleiotropy, in which the same genetic variants independently influence multiple CVDs. Formally distinguishing these models would require family- or twin-based designs not yet systematically applied to CVD comorbidity (101).

Replication analyses in East Asian ancestry cohorts provided complementary support for shared genetic architecture across populations. For CVDs with adequate power (AAA, AF, IHD, HF), heritability estimates and cross-trait genetic correlations were comparable between EUR and EAS ancestries, in line with previous reports of comparable common-variant effects (102,103). Although limited power restricted replication to a subset of traits, several pleiotropic associations showed directional consistency, and mediation by key risk factors such as smoking and type 2 diabetes was observed, underscoring the broad relevance of shared cardiovascular mechanisms across populations.

Several limitations merit consideration. Differences in GWAS power across traits and ancestries inherently limit detection of shared signals and direct comparison of overlap between ancestries, particularly for less-powered traits and non-European populations. Moreover, common genetic variation explained only a modest proportion of phenotypic comorbidity, indicating that environmental exposures, clinical factors, and rare variants contribute substantially to CVD co-occurrence beyond the scope of this study. Finally, mediation analyses reflect shared genetic covariance rather than causal directionality. Phenotypic associations may also be attenuated by the healthier-than-average composition of UK Biobank participants.

In conclusion, our findings indicate that cardiovascular comorbidity is driven by a combination of systemic risk factors, extensive pleiotropy across cardiovascular conditions, and shared biological processes centred on the vascular wall, particularly across arterial, valvular, and arrhythmia-related phenotypes. Integration of gene-, cell-type-, and tissue-level evidence highlights endothelial and mural cell populations as a common substrate linking diverse cardiovascular phenotypes, supported by prioritised genes such as *SMAD6*, *QKI*, *NME7*, and *PKD1.* These results emphasize the value of cross-disease genetic approaches for dissecting shared cardiovascular biology and suggest that targeting risk factors with widespread genetic influence across CVDs—such as adiposity-related traits and metabolic dysregulation identified in our mediation analyses—may offer effective strategies to reduce cardiovascular multimorbidity.

## Acknowledgements

C.R., D.W., M.S., D.P. and S.v.d.S were funded by NWO Gravitation: BRAINSCAPES: A Roadmap from Neurogenetics to Neurobiology (grant no. 024.004.012 [to D.P.]). D.P. was funded by a European Research Council advanced grant (no. ERC-2018-AdG GWAS2FUNC 834057 [to D.P.]). EvW is financially supported by funds of the Academic Education and Research sector plans of the Dutch Ministry of Education, Culture and Science. S.J.J. received research support from the Dutch Heart Foundation (03-007-2022-0035), the Amsterdam UMC, and the KIC program through the Dutch Research Council and Dutch Heart Foundation (PRECISE project). C.R.B. was supported by funding from the Dutch Heart Foundation (CVON 2018-30 PREDICT2), the Pathfinder Cardiogenomics programme of the European Innovation Council of the European Union (DCM-NEXT and NaV1.5-CARED projects), and the KIC program through the Dutch Research Council and Dutch Heart Foundation (PRECISE project).

This work was carried out on the Genetic Cluster Computer, which is financed by the Netherlands Scientific Organization (NWO: 480-05-003), by the VU University, Amsterdam, the Netherlands, and by the Dutch Brain Foundation, and is hosted by the Dutch National Computing and Networking Services SurfSARA. This research has been conducted using the UK Biobank resource under application numbers 16406 and 176602. We thank the numerous participants, researchers, and staff from many studies who collected and contributed to the data. In particular, we gratefully acknowledge the contributions of the All of Us and UK Biobank participants, whose involvement made this research possible. We also thank the National Institutes of Health’s All of Us Research Program for granting access to the participant data utilized in this study. Graphical abstract, Figure 1 and Figure 6 were Created in BioRender with license to publish.

## Author Contributions

C.R.: Methodology, Formal Analysis, Writing, Visualisation, Project Administration. D.W.: Methodology, Formal Analysis, Writing. E.v.W.: Methodology, Feedback. S.J.J.: Resources, Methodology, Feedback. M.E.C.: Resources, Feedback. P.H.: Resources, Feedback. C.B.: Methodology, Feedback M.S.: Formal Analysis, Feedback. D.P.: Feedback, Methodology, Funding Acquisition, Resources. S.vd.S.: Conceptualisation, Supervision, Feedback, Methodology, Writing.

## Data availability statement

GWAS and ExWAS summary statistics of all studied CVD traits will upon publication become available at https://ctg.cncr.nl/software/summary_statistics.

All GWAS summary statistics are based on Human Genome Build 37 (GRCh37/hg19). Precomputed EUR and EAS LD scores and HapMap 3 reference file were obtained from: <u>Link</u> 1000 Genomes Phase 3 Project EUR and EAS LD reference data: <u>Link</u> In-sample LD reference from UKB: https://broad-alkesgroup-ukbb-ld.s3.amazonaws.com/UKBB_LD/

In-sample LD reference from FinnGen Google Cloud storage available upon access from here: https://elomake.helsinki.fi/lomakkeet/124935/lomake.html

Gene information was obtained from the GeneCard database (v5.12.0 Build 702; https://www.genecards.org).

SNP information was obtained from Open Target Genetics: https://genetics.opentargets.org

## Software Availability

FLAMES(v1.1.0) https://github.com/Marijn-Schipper/FLAMES

LAVA(v0.1.0) local genetic correlations: https://ctg.cncr.nl/software/lava

Regenie(v3.4.13): https://github.com/rgcgithub/regenie

