## Supplementary Notes and Figures for "Mapping the genetic landscape of eight common cardiovascular diseases"

### Supplementary Notes

#### Supplementary Note 1. Extended phenotype definition and description

After selecting the phenotypes within UK Biobank (UKB) and defining them with the appropriate ICD-10 codes, we searched the FinnGen database for publicly available GWAS with similar phenotype definitions. FinnGen GWAS summary statistics were harmonized by lifting over the SNP genomic positions from genome build 38 to genome build 37 (GRCh37/hg19).

In the UKB, patient disease occurrences were derived from ICD-10 subfields within data category 1712 ("First occurrences"), which includes primary care data (category 3000), hospital inpatient data (category 2000), Death Register records (fields 40001-40002), and self-reported medical conditions (field 20002) reported at the first or subsequent visits to UKB assessment centers. All UKB CVD GWAS used the same controls (N = 342,432) consisting of people without any diagnosis of the 8 analysed CVDs. FinnGen applied phenotype-specific control exclusions: atrial fibrillation excluded controls with other cardiovascular conditions (hypertension, ischaemic heart disease, pulmonary embolism, cerebrovascular disease, aortic aneurysm, venous/lymphatic disease, and other heart diseases); abdominal aortic aneurysm excluded controls with other aortic aneurysm diagnoses (I9_DOAAC); heart failure excluded controls with non-specific heart failure (I9_HEARTFAIL_NS); varicose veins excluded controls with other venous or lymphatic disorders (I9_DISVEINLYMPH); and conduction disorders excluded controls with other heart diseases (I9_OTHHEART). An overview of the included CVD definitions from both FinnGen and UKB is provided below:

*Abdominal Aortic Aneurysm (AAA)*

In the UKB GWAS, cases for AAA were identified based on ICD-10 codes I71.3 (Abdominal aortic aneurysm, ruptured) and I71.4 (Abdominal aortic aneurysm, without mention of rupture). The same case definition was used within FinnGen. In UKB, a total of 2,769 cases of AAA were identified, while 3,869 cases were diagnosed within FinnGen. The genetic correlation between the GWAS summary statistics of these two cohorts was high (*r*_g_ = 0.99, P = 2.71 x 10^-6^).

*Atrial Fibrillation (AF)*

In the UKB GWAS, AF cases were identified using ICD-10 code I48 (Atrial fibrillation and flutter). The same definition was used in FinnGen. A total of 39,080 AF cases were diagnosed in the UKB and 50,743 cases in FinnGen. The GWAS summary statistics of these 2 cohorts showed a high genetic correlation (*r*_g_ = 0.87, P = 5.15 x 10^-203^).

*Aortic Valve Stenosis (AS)*

Cases for AS in the UKB GWAS were identified using ICD-10 codes I35.0 (Aortic (valve) stenosis) and I35.2 (Aortic (valve) stenosis with insufficiency). The same case definition was applied in FinnGen. In the UKB, 5,820 AS cases were identified, and 9,870 cases were diagnosed in FinnGen. The GWAS summary statistics of these 2 cohorts showed a high genetic correlation (*r*_g_ = 0.84, P = 1.90 x 10^-16^).

*Conduction Disorders (CD)*

CD cases in the UKB GWAS were identified using ICD-10 codes I44 (Atrioventricular and left bundle-branch block) and I45 (Other conduction disorders). The same definition was used in FinnGen. In the UKB, 18,689 CD cases were identified, while 10,944 cases were diagnosed in FinnGen. The GWAS summary statistics of these 2 cohorts showed a high genetic correlation (*r*_g_ = 0.76, P = 1.19 x 10^-7^).

*Heart Failure (HF)*

In the UKB GWAS, HF cases were identified using ICD-10 code I50 (Heart failure). The same definition was used in FinnGen. In the UKB, 18,366 HF cases were identified, while 29,672 cases were diagnosed in FinnGen. The GWAS summary statistics of these 2 cohorts showed a high genetic correlation (*r*_g_ = 0.85, P = 7.53 x 10^-20^).

*Ischemic Heart Disease (IHD)*

IHD cases in the UKB GWAS were identified using ICD-10 codes I21 (Acute myocardial infarction), I22 (Subsequent myocardial infarction), I24 (Other acute ischaemic heart diseases), and I25 (Chronic ischaemic heart disease). In FinnGen, the same codes were used, along with I20 (Angina pectoris) and I23 (Certain current complications following acute myocardial infarction). In the UKB, 52,886 IHD cases were identified, and 69,008 cases were diagnosed in FinnGen. The GWAS summary statistics of these 2 cohorts showed a high genetic correlation (*r*_g_ = 0.91, P = 1.25 x 10^-142^).

*Venous Thromboembolism (VTE)*

VTE cases in the UKB GWAS were identified using ICD-10 codes I26 (Pulmonary embolism), I80.1 (Phlebitis and thrombophlebitis of femoral vein), I80.2 (Phlebitis and thrombophlebitis of other deep vessels of lower extremities), and I82.2 (Embolism and thrombosis of vena cava). In FinnGen, the definition included I26, I80 (Phlebitis and thrombophlebitis), and O87.1 (Deep phlebothrombosis in the puerperium). In the UKB, 13,189 VTE cases were identified, while 21,021 cases were diagnosed in FinnGen. The GWAS summary statistics of these 2 cohorts showed a high genetic correlation (*r*_g_ = 0.89, P = 1.42 x 10^-22^).

*Varicose Veins (VV)*

VV cases in the UKB GWAS were identified using ICD-10 code I83 (Varicose veins of lower extremities). The same definition was used in FinnGen. In the UKB, 23,443 VV cases were identified, while 31,719 cases were diagnosed in FinnGen. The GWAS summary statistics of these 2 cohorts showed a high genetic correlation (*r*_g_ = 0.85, P = 7.23 x 10^-127^).

#### Supplementary Note 2. GWAS and case-control definition and study design

To investigate the shared genetic underpinnings of the cardiovascular diseases (CVDs) included in our study, we conducted genome-wide association studies (GWAS) using data from individuals with diagnosed CVDs reported in the UKB. Our primary goal was to identify shared, or pleiotropic, genetic effects between these CVDs. We sought to address two issues that might confound results related to pleiotropy:

1	*GWAS-case overlap*: Patients with multiple diagnosed CVDs (comorbidities) can appear in GWAS analyses for more than one disease. This case-overlap between GWAS can complicate the interpretation of genetic overlap analyses, as it becomes challenging to discern if genetic similarities between diseases are due to actual pleiotropy or simply the result of overlapping sample populations.

2	*GWAS-control definitions*: The definition of controls can influence the genetic correlations observed between diseases.

*GWAS-case overlap*

When conducting disease-specific GWAS with the aim of assessing genetic overlap between CVDs, individuals with multiple diagnoses may be included as cases in more than one GWAS, leading to overlap of cases across analyses. We therefore considered case definitions:

•	*Lenient definition*: This approach includes all patients diagnosed with a particular CVD, regardless of comorbidities, resulting in case-overlap between CVD GWAS. While this maximizes the sample size for each GWAS, it does not account for overlapping samples in downstream analyses. Thus, it risks interpreting results as pleiotropic that, in reality, are due to comparing allele frequencies of the same individuals.

•	*Stringent definition*: This approach excludes all comorbid cases, analysing only those patients diagnosed with a single CVD. While this eliminates case overlap and makes interpretation of pleiotropy more straight-forward, it also reduces sample size and statistical power. Additionally, excluding comorbid cases may not accurately represent diseases with high levels of comorbidity, potentially skewing the results toward a specific, possibly genetically unique, subgroup of the disease.

•	*No-case-overlap definition*: As a compromise, we randomly assigned patients with comorbidities to only one of their diagnosed CVDs and excluded them from analyses of other CVDs. This method avoids case overlap between CVD GWAS while utilizing all comorbid cases to increase GWAS power. Our main analyses focused on results from this no-case-overlap approach. As a validation step, we conducted genetic correlations among CVD-pairs based on each of the above three definitions, separately. We observed substantial inflation of *P*-values under the lenient definition, such that all CVD pairs were identified as significantly genetically correlated (**Supplementary Figure 1**). Compared with the no-overlap definition, *P*-values from the lenient definition had a median of 48 billion times more significant estimation. In addition, the genetic covariance intercept showed a very strong correlation with the Z-scores of the genetic correlations (*r* = 0.93), indicating that the observed significance was largely driven by shared sample structure and overlap rather than by true increases in underlying genetic signal.

*Control Definitions*

The control group for each UKB-based CVD GWAS comprised individuals with no diagnosis for any of the eight studied CVDs, totalling 342,432 people. Using the same control group across all GWAS might introduce genetic overlap between CVDs due to specific enrichments for allele frequencies in the control group. To address this potential bias, we used FinnGen as a replication cohort to verify that pleiotropic effects observed within UKB were also observed to be pleiotropic within FinnGen.

*Study Design*

Our study design aims to navigate the challenges of case overlap and control definitions on the one hand, and statistical power/optimal use of data on the other. By implementing the no-case-overlap definition (**Supplementary Figure 2**) and using a discovery-replication study design, we seek to accurately identify shared genetic effects among common CVDs that are not inflated due to GWAS case-control definitions.

#### Supplementary Note 3. Handling close relatives and X chromosome

GWAS were conducted using UKB data to investigate the genetic underpinnings of the 8 CVD phenotypes. The analyses were performed using REGENIE v3.4.13, which accounts for genetic relatedness among individuals. Covariates included in the GWAS model were age, sex, genotyping array, and the first 20 principal components (PCs) to account for population stratification.

To address induced genetic overlap caused by familial relationships between GWAS cases, we identified any third-degree relatives or closer (N_total relative-pairs_ = 8,541) from the GWAS case groups to ensure independent sampling. If detected relatives were diagnosed with the same CVD (N_relatives_ = 3,954), they were retained in the analysis as cases for that CVD. Their genetic relatedness was then controlled within the GWAS analysis using REGENIE. If relatives had different CVD diagnoses, one was randomly excluded from the analysis to prevent induced genetic similarity across CVD GWASs (N_excluded_ = 4,587).

REGENIE GWAS analyses were performed in two steps: 1) REGENIE estimates a whole-genome regression model using ridge regression on a subset of genetic variants to capture genetic relationships among individuals, employing a leave-one-chromosome-out (LOCO) approach to avoid biases. This model predicts individual-level genetic effects, which account for population structure and relatedness. 2) The predicted genetic effects from step 1 are used as covariates in single-variant association tests for each genetic variant. Association analyses incorporated the use of Firth regression that addresses sample size inequalities by applying Firth’s penalized likelihood method to provide more accurate genetic effect estimates in the presence of imbalanced case-control ratios (i.e., many more controls than cases).

The GWAS included the X chromosome and its pseudo-autosomal regions (PARs). The PARs were treated as autosomes, while males were coded as diploids for variants in non-PAR X regions, assuming random X-inactivation. Heterozygous male calls were set as missing, and missingness filters were reapplied. This resulted in a total of 267,924 variants on the X chromosome after filtering for minor allele frequency (MAF > 0.01).

#### Supplementary Note 4. Extended MAGMA methods

*Gene-based association analysis*

Gene-based genome-wide association analyses were performed using MAGMA, which aggregates SNP-level association statistics into gene-level tests while accounting for LD. SNPs were mapped to protein-coding genes based on GRCh37 (hg19) coordinates, yielding 19,427 genes. Gene-based association analyses were conducted separately for each CVD GWAS.

For discovery analyses in UKB, multiple testing across genes was controlled using FDR correction at 5%. Genes reaching FDR significance for a given CVD were evaluated for replication in the corresponding FinnGen GWAS using Bonferroni correction based on the number of UKB-significant genes for that phenotype.

Genes replicated in at least one CVD were subsequently tested for pleiotropic association across all UKB-based CVDs using FDR correction (5%). Pleiotropic genes were finally assessed for replication across FinnGen CVDs using Bonferroni correction based on the number of pleiotropic candidates.

*Gene-set enrichment analysis*

Gene-set analyses were conducted using curated gene sets from MSigDB (v2023.1.Hs). MAGMA gene-set tests assessed whether genes with stronger association signals were overrepresented within predefined biological pathways. The same four-stage framework was applied as for gene-based analyses:

- Discovery of significant gene sets within each UKB-based CVD using FDR < 5%.
- Replication of UKB-significant gene sets in the corresponding FinnGen CVD using Bonferroni correction.
- Pleiotropy testing of replicated gene sets across UKB-based CVDs using FDR < 5%.
- Replication of pleiotropic gene sets across FinnGen CVDs using Bonferroni correction.

*Tissue- and cell-type gene-property analyses*

Tissue- and cell-type enrichment analyses were performed using MAGMA gene-property models, which test whether gene-level association statistics are associated with gene expression profiles. Bulk tissue expression data were obtained from GTEx v8 (54 tissues). Cell-type expression profiles were derived from GTEx single-cell annotations (500 cell types). For each annotation, MAGMA tested whether higher relative expression predicted stronger gene-level associations, adjusting for average expression across annotations. One-sided tests were used to identify annotations showing enrichment. For both tissue- and cell-type analyses, the same four-stage framework was applied:

- UKB discovery using FDR < 5% across annotations.
- Replication in FinnGen using Bonferroni correction.
- Pleiotropy testing across UKB-based CVDs using FDR < 5%.
- Replication of pleiotropic annotations across FinnGen CVDs using Bonferroni correction.

*Rare-variant gene-set and gene-property analyses*

Rare-variant enrichment analyses were performed using gene-level pLOF burden statistics from UKB and Cauchy-combined burden results from the All of Us cohort. Gene-level p-values were converted into MAGMA-compatible Z-scores using the inverse normal transformation (qnorm in R), assuming negligible LD between genes. The number of variants mapped per gene was matched to UKB where possible; genes absent in UKB were assigned a count of one variant.

Gene-set analyses were conducted using MAGMA v1.10 across the full MSigDB v2023.2.Hs collection. Tissue-property analyses were performed using GTEx v7 bulk tissue expression profiles (54 tissues), testing whether gene expression predicted rare-variant burden Z-scores while adjusting for average expression across tissues. Rare-variant gene-set and gene-property analyses followed the same 4-step framework of discovery, replication, pleiotropy, and replication-of-pleiotropy used for common-variant analyses.

#### Supplementary Note 5. Extended rare variant gene burden test

Genetic variants from whole-genome sequencing data in both UKB and All of Us were annotated using VEP (v105) with the LOFTEE plugin, the dbNSFP database (v4.2), PrimateAI-3D, popEVE, and AlphaMissense (1–5). Annotations were restricted to ENSEMBL canonical transcripts of protein-coding genes. Missense variants were defined according to VEP annotation, and high-confidence protein-truncating variants (PTVs) were identified using LOFTEE. A stringent PTV subset (PTVnoflag) excluded any variants carrying LOFTEE flags. Ancestry-specific allele frequencies from gnomAD v2 were used to derive the maximum population minor allele frequency (MAF_MAX_) across non-Finnish European, African, South Asian, East Asian, and Admixed American super-populations.

We defined the following variant masks: (1) PTV, all high-confidence PTVs regardless of LOFTEE flags; (2) PTVnoflag, PTVs without LOFTEE flags; (3) missense variants predicted damaging by PrimateAI-3D; (4) missense variants predicted damaging by at least 3 of 4 tools (PrimateAI-3D, REVEL, popEVE, AlphaMissense) (6); and (5–6) PTV+missense combinations of masks 1–2 with masks 3–4. Each mask was evaluated at MAF thresholds of <0.01, <0.001, and <0.00001 based on both in-sample allele frequency and MAF_MAX_.

Burden analyses were performed using REGENIE v4.1 (7), following its standard two-step framework. In step 1, individual-level genetic predictions (null models) were generated using high-quality common variants genome-wide (from DRAGEN WGS files in UKB or ACAF files in All of Us) after variant QC and LD pruning, using the --loocv, --bt, and --write-null-firth flags. Separate null models were fit for each cardiovascular phenotype. In step 2, gene-level burden tests were performed using exome sequencing data and the variant masks defined above, using approximate Firth's regression with back-correction of standard errors. In UKB, masks were constructed using REGENIE's default --build-mask max rule under an additive genetic model. In All of Us, a dominant burden model was applied. All analyses were adjusted for the first 20 principal components, age, age², and genetically inferred sex. UKB analyses were additionally adjusted for assessment region, genotyping batch, and availability of GP records.

To obtain a single gene-level association statistic integrating evidence across all burden masks, we applied the Cauchy combination test (CCT). Masks were restricted to variants affecting ENSEMBL canonical transcripts with a cumulative minor allele count (cMAC) > 20; singleton-only masks were excluded. For each gene, p-values from qualifying masks were combined using equal weights to derive an overall Cauchy p-value. A separate rare-variant-specific Cauchy p-value was computed by restricting the combination to masks with MAF < 0.001. Genes lacking any qualifying rare masks were assigned missing values for the rare-specific statistic. Genes were ranked by their overall Cauchy p-value in the final output.

To identify genes with pleiotropic rare-variant associations across CVDs, we used a two-stage replication strategy. First, within each CVD in UKB, genes reaching FDR < 5% (Benjamini–Hochberg) based on their overall Cauchy p-value were selected as significant. Second, these gene-level associations were evaluated for independent replication in All of Us rare-variant summary statistics, with replication defined as P < 0.05 after Bonferroni correction for the number of genes tested. Genes that replicated in All of Us were subsequently tested for pleiotropic associations across all CVDs in UKB (FDR < 5% within the set of replicated genes), and pleiotropic gene–phenotype pairs were further evaluated in All of Us using Bonferroni correction for the number of pleiotropic associations tested.

#### Supplementary Note 6. Univariate results from GWAS and ExWAS

We performed common variant GWAS analyses for all 8 CVDs in the UKB and used these summary statistics to assess the genetic overlap between CVDs. We replicated the findings from the UKB using GWAS summary statistics from Finngen. The effective sample sizes of these traits ranged from 5754 (AAA) to 127,353 (IHD) in the UKB and from 14,554 (AAA) to 218,327 (IHD) in Finngen (**Supplementary Table 1**). The SNP heritability estimates in the UKB were relatively similar to the estimates from Finngen with the exception of AS which was almost double in the UKB Biobank (21% (SE = .036) vs 12% (SE = .014); **Supplementary Table 2**). Overall, the sample size and SNP heritability of these traits were sufficiently large to captured enough association signal for follow-up comparisons. FUMA identified a limited number of independent lead SNPs for AAA, AS, CD, and HF (2–5 loci), whereas substantially more loci were detected for AF (83), IHD (63), VTE (21), and VV (53). Gene-based analysis using MAGMA showed a similar pattern, with few or no significant genes for AAA, AS, CD, and HF (0–3 genes), but a larger number of associated genes for AF (127), IHD (67), VTE (16), and VV (75).

We performed rare variant (MAF<1%) pLOF burden analysis across 19,549 protein coding genes in the UKB for all 8 CVDs to identify overlap in genes and gene-sets associated with the CVDs. We replicated findings from the UKB using the All of Us mixed ancestry rare variant (MAF<0.1%) pLOF and missense burden analysis summary statistics from Jurgens et al. (2024) (8)) (18,016 genes). The same set of UKB individuals included in the common variant analyses were included in the rare variant analyses with the effective sample size ranging from 5825 (AAA) to 129,168 (IHD). The replication dataset (All of Us) was similar in size with an effective sample size ranging from 743 (AS) to 166,554 (CD). The 95% genomic inflation factors ranged from 0.93 (AS) to 1.021 (VTE) in the UKB and 1.005 (Bundle branch block (CD)) to 1.10 (Aortic valve disease (AS)) in All of Us (9). The number of FDR-BH <5% genes range from 0 to 6 (AF) in the UKB and 0 to 2 (multiple) in All of Us. Overall, the rare variant association analyses did not identify as much association signal as the common variant analyses but there was still sufficient signal to identify significant genes and gene-sets.

#### Supplementary Note 7. Sex-stratified GWAS of CVDs in UKB

Sex-stratified GWAS were conducted for eight CVDs in UKB to assess potential differences in genetic liability between males and females. For each CVD, GWAS were performed separately in males and females using identical analytical pipelines. To minimize confounding due to differential statistical power, sex-stratified GWAS were down-sampled to achieve matched case–control ratios between males and females within each phenotype. The resulting effective sample sizes (Neff = 4 x N_cases x N_controls / (N_cases + N_controls)) were the therefore the same for males and females at: AAA, Neff = 902; AF, Neff = 35,735; AS, Neff = 4,185; CD, Neff = 15,204; IHD, Neff = 43,274; HF, Neff = 11,902; VTE, Neff = 17,040; and VV, Neff = 24,519.

Cross-sex LDSC analyses indicated largely shared genetic architectures between males and females across all CVDs. Genetic correlation estimates were generally close to unity, with confidence intervals overlapping *r*_g_ = 1 for all traits (**Supplementary Table 1**). The cross-sex *r*_g_ for VTE showed the widest confidence interval and marginally included unity, possibly indicating that higher-powered sex-stratified GWAS may be required to resolve potential differences in genetic liability for this phenotype.

SNP-level sex-heterogeneity tests did not identify any genome-wide significant loci. However, across all CVDs, multiple loci showed suggestive evidence of sex-dependent effects (5 × 10⁻⁸ < *P* < 1 × 10⁻⁵), including AAA (9 loci), AF (14 loci), AS (11 loci), CD (12 loci), HF (5 loci), IHD (10 loci), VTE (11 loci), and VV (11 loci), suggesting that sex-dependent effects might be uncovered with higher powered sex-stratified GWAS.

Overall, sex-stratified analyses did not provide strong evidence for widespread sex-specific genetic architectures across CVDs but future analyses with larger sample sizes are required for more confident conclusions.

#### Supplementary Note 8. Shared genes from loci where FLAMES predictions were not resolved

Across the 13 loci with evidence for shared genes between CVD pairs, 5 loci could not be unambiguously resolved by automated gene prioritization for various reasons, including genes being non-coding, complex linkage disequilibrium and high gene density, rare causal variants, or discordance between statistical prioritisation and biological plausibility.

The clearest example is the Chr.9p21.3 region shared between IHD and AAA, where the well-established causal signal resides in the non-coding transcript CDKN2B-AS1 (10–16). This gene could not be prioritised by FLAMES due to non-coding genes not being included in the analysis. Similarly, at Chr.19p13.2, *LDLR* was prioritised for AAA, whereas IHD implicated the adjacent gene *SMARCA4*; however, the locus is gene-dense with complex LD, and *LDLR* influence on IHD is supported by the ExWAS analyses in addition to extensive functional literature (17–19), making *LDLR* the likely shared gene in this locus. At Chr.6q25.3–q26, AS and IHD prioritised *SLC22A3*, while AAA prioritised *LPA*. Although *SLC22A3* may contribute to disease risk, *LPA* represents the most plausible causal gene across all three of these disorders, consistent with the strong and well-established role of lipoprotein(a) in atherosclerotic and valvular disease and the known difficulty of tagging *LPA* kringle variation with common SNPs (20–25). At the Chr.1q24.2 locus, *NME7* was prioritised for both VTE and IHD; however, the nearby gene *F5* is a canonical VTE risk gene (26), and the causal variant (Factor V Leiden) is rare and not present in the CVD GWAS panels. Consequently, *F5* is likely the main driver of the genetic signal in VTE and previous studies have suggested that the same causal variant linked to *F5* confers IHD risk (27) and other CVD comorbidities as well (28). Likewise, at the Chr.14q12 locus where *AKAP6* was prioritised for AF and *ARHGAP5* for CD, *AKAP6* represents the more biologically plausible effector gene given its established role in calcium signalling and cardiac muscle contraction (29,30) and the lead SNP associations reside for both traits within the *AKAP6* gene (see **Supplementary Figures** **5-17** for all locuszoom plots).

#### Supplementary Note 9. Local genetic mediation analysis using risk factor traits

To investigate whether significant local genetic correlations between CVDs could be explained by shared genetic covariance with other traits, we performed conditional local genetic correlation analyses using LAVA. This framework tests whether the marginal local genetic correlation between two CVDs is attenuated after conditioning on a third covariate trait, indicating potential genetic mediation.

GWAS summary statistics for 288 potential covariate traits were considered, including blood biomarkers, established cardiovascular risk factors, and potentially confounding traits such as BMI, social deprivation, and smoking initiation. Prior to conditional analyses, covariates were required to show a significant local univariate genetic signal (*P* < 1 × 10⁻⁴) and nominal genetic correlation with both CVDs in at least one of the CVD pairs. After filtering, 80 covariate traits remained, covering 14 CVD pairs across 25 genomic loci.

Conditional analyses CVDs identified 18 significant covariate effects after correction for the number of CVD pairs tested (*P* < 3.57 × 10⁻³). Of these, nine effects involved serum cholesterol–related traits (LDL, ApoB, Lp(a)) within a single genomic region on chromosome 6 (160.6–161.4 Mb), shared between AS, AF, and IHD. After correcting for all conditional tests performed, only erythrocyte distribution width remained significant as a mediator, specifically for the VTE–VV pair within a locus on chromosome 9 (136.0–136.8 Mb; **Supplementary Figure 19**).

Replication across datasets was limited: only the conditional effect of serum ApoB in the chromosome 6 region between AF and IHD showed nominal significance in both UKB- and FinnGen-based analyses, indicating that most observed mediation effects should be interpreted cautiously.

Follow-up conditional analyses in correlated genomic regions further supported a partial contribution of serum cholesterol–related traits to the shared genetic signal between AS, AF, and IHD within the chromosome 6 locus (Supplementary Note 2; Supplementary Table 13). Overall, these analyses suggest that local genetic correlations between certain CVD pairs may, in specific regions, be partially attributable to shared genetic influences on cardiometabolic risk factors, although robust mediation effects were limited.

#### Supplementary note 10. Description of pairwise CVDs with some shared genetic signal

Only trait that showed any genetic overlap in terms of genetic correlations, genes, annotations or shared risk factors are described below.

*Heart failure - ischaemic heart disease:*

Heart failure and ischemic heart disease showed the strongest genome-wide overlap, with a global genetic correlation of ***r*_g_ = 0.66 (SE = 0.06)**. The local genetic correlation analysis identified **four** shared regions (loci **1398, 1852, 2233, 970**), indicating that shared common-variant signal arises from multiple genomic blocks. Gene-level evidence converged across variant classes: FLAMES prioritised shared genes including *LDLR* and *TTN*, and rare-variant analyses additionally implicated *PKD1*. These findings point to shared mechanisms spanning lipid metabolism and myocardial structural biology. This pair also showed the largest number of significant blood-metabolite attenuation effects, consistent with partial mediation through circulating metabolic and lipid-related pathways.

*Abdominal aortic aneurysm – ischaemic heart disease:*

Abdominal aortic aneurysm and ischemic heart disease showed substantial overlap with ***r*_g_ = 0.51 (SE = 0.11)**. Locus-level evidence was prominent, with **three** local ***r*_g_** regions (loci **1398, 1722, 2318**) and **seven** colocalising signals (loci **1, 49, 53, 56, 70, 76, 89**). FLAMES prioritised **six** shared genes (***CDKN2B-AS1, CELSR2, LPA, SLC22A3, LDLR, ZNF259***), implicating arterial-wall biology and lipid pathways. These findings align with extensive prior evidence linking 9p21 (***CDKN2B-AS1)*** and lipoprotein pathways to both aneurysmal and atherosclerotic disease.

*Atrial fibrillation–heart failure:*

Atrial fibrillation and heart failure had a large global genetic correlation (***r*_g_ = 0.55, SE = 0.06**) and clustered within the same latent genetic factor (**F1, electromechanical**). The local genetic correlation analysis identified **four** shared regions (loci **1037, 113, 1271, 919**), and colocalisation supported **two** shared loci (**17, 52**). FLAMES prioritised CPEB4 and PITX2, while rare-variant analyses implicated TTN, LMNA, and PKD1. Gene-set enrichments highlighted myocardial structural and conduction-related processes, including fibrosis-related pathways, consistent with shared myocardial remodelling and electrophysiological mechanisms.

*Conduction disorders–heart failure:*

Conduction disorders and heart failure showed strong genome-wide overlap (***r*_g_ = 0.63, SE = 0.13**) and loaded on the same gSEM factor (**F1, electromechanical**). The local genetic correlation analysis identified **two** shared regions (loci **1743, 970**). Although no shared FLAMES genes were prioritised, rare-variant analyses implicated *TTN* and *LMNA*, supporting shared myocardial structural and conduction-related processes.

*Conduction disorders–ischaemic heart disease:*

Conduction disorders and ischemic heart disease had a large genome-wide overlap (***r*_g_ = 0.61, SE = 0.10**). The local genetic correlation analysis identified **two** shared regions (loci **1613, 970**. No shared genes were prioritised, however the gene set *left anterior fascicular block* was significant for both based on rare variants. Genetic mediation analyses implicated adiposity related traits (BMI, WHR) and hypertension.

*Aortic stenosis–ischaemic heart disease:*

Aortic stenosis and ischemic heart disease showed substantial overlap (***r*_g_ = 0.43, SE = 0.07)** and clustered in the same latent factor (**F3, obstructive**). The local genetic correlation analysis identified **two** shared regions (loci **1084, 2079**), and colocalisation supported **three** shared loci (**49, 56, 70**). FLAMES prioritised **three** shared genes (***CELSR2, LPA, SLC22A3***), implicating a lipoprotein-related region that is well supported by prior genetic and translational evidence. In particular, extensive literature links **LPA / lipoprotein(a)** to both coronary outcomes and calcific aortic valve stenosis, consistent with the shared-locus pattern observed here.

*Atrial fibrillation–ischaemic heart disease:*

Atrial fibrillation and ischemic heart disease showed moderate genome-wide overlap (***r*_g_ = 0.29, SE = 0.03**). The local genetic correlation analysis identified **five** shared regions (loci **1084, 1087, 2043, 2072, 698**). FLAMES prioritised **two** shared genes (***QKI*** and ***TLE3***). ***QKI*** has published links to vascular smooth muscle cell phenotypic regulation and endothelial biology, providing a plausible cellular context for shared AF–IHD risk in analyses that also implicate vascular support and stromal programs across CVDs.  ***TLE3*** has established roles in metabolic and adipocyte differentiation programs, consistent with broader findings in which adiposity-related traits frequently attenuate shared CVD genetic covariance in conditioning analyses. Rare-variant analyses implicated *TTN* and *PKD1*. Gene-set enrichments shared between AF and IHD were concentrated in arrhythmia/conduction-related gene-sets.

*Atrial fibrillation–Conduction disorder:*

Atrial fibrillation and conduction disorders showed substantial sharing (***r*_g_ = 0.46, SE = 0.10**) and clustered within **F1 (electromechanical)**. One shared local ***r*_g_** region was detected (locus **1965**), and one colocalising locus was observed (**11**). FLAMES prioritised *AKAP6* and *TTN*, and rare-variant analyses additionally implicated *LMNA*. The gene set *left anterior fascicular block* was significant for both based on rare variants. This convergence supports shared myocardial structural and electrophysiological mechanisms.

*Ischemic heart disease–venous thromboembolism:*

Ischemic heart disease and venous thromboembolism showed clear genome-wide overlap (***r*_g_ = 0.39, SE = 0.05**). One shared local ***r*_g_** region was detected (locus **1479**), and one colocalising locus was observed (**91**). FLAMES prioritised *NME7/F5*, and rare-variant analyses implicated *PKD1*. Mediation analyses identified multiple shared risk factors, including adiposity and alcohol-related traits, supporting shared systemic liability.

*Venous thromboembolism–varicose veins:*

Venous thromboembolism and varicose veins showed moderate overlap (***r*_g_ = 0.35, SE = 0.05**) and clustered within **F2 (vascular integrity)**. Two shared local regions and two colocalising loci were identified, but no shared genes were prioritised, indicating overlap at the regional level.

*Abdominal aortic aneurysm–venous thromboembolism:*

Abdominal aortic aneurysm and venous thromboembolism showed moderate-to-large overlap (***r*_g_ = 0.42, SE = 0.15**) and clustered within **F2 (vascular integrity)**. No shared loci or genes were identified, but genetic mediation analyses implicated BMI, suggesting shared cardiometabolic risk.

*Abdominal aortic aneurysm–varicose veins:*

Abdominal aortic aneurysm and varicose veins showed moderate overlap (***r*_g_ = 0.23, SE = 0.11**) and clustered within **F2 (vascular integrity)**, without additional supporting evidence at locus or gene level.

*Heart failure–venous thromboembolism:*

Heart failure and venous thromboembolism showed moderate overlap (***r*_g_ = 0.39, SE = 0.09**). Although no shared loci were identified, rare-variant analyses implicated *PKD1*, and mediation analyses highlighted adiposity and alcohol-related traits, suggesting shared systemic risk pathways.

*Atrial fibrillation–venous thromboembolism:*

Atrial fibrillation and venous thromboembolism showed moderate overlap (***r*_g_ = 0.23, SE = 0.06**). No shared loci were identified, but rare-variant analyses implicated *PKD1*, and mediation analyses indicated contributions from cardiometabolic and behavioural risk factors.

*Ischemic heart disease–varicose veins:*

Ischemic heart disease and varicose veins showed modest overlap (***r*_g_ = 0.15, SE = 0.04**), but had **three** colocalising loci (**108, 116, 94**). No shared genes were prioritised, but enrichment analyses implicated vascular cell types (*endothelial, pericytes, fibroblasts*) and *coronary artery tissue*, supporting shared vascular biology.

*Atrial fibrillation–varicose veins:*

Atrial fibrillation and varicose veins showed a small but non-zero overlap (***r*_g_ = 0.10, SE = 0.04**). Two shared local ***r*_g_** regions were detected (loci **2098, 2168**), and FLAMES prioritised **one** shared gene (***IGF1R***). Functional enrichment analyses supported involvement of vascular and stromal cell types (*endothelial, pericytes, fibroblasts*). *IGF1R* signalling has been linked in experimental and translational work to myocardial remodelling and fibrosis-related processes, providing a plausible mechanism through which shared variants might influence both rhythm-related phenotypes and vascular traits, although the interpretation remains hypothesis-generating.

*Aortic stenosis–heart failure:*

Aortic stenosis and heart failure showed moderate overlap (***r*_g_ = 0.28, SE 0.10**). No shared loci were identified, but rare-variant analyses implicated *LDLR*, and mediation analyses highlighted BMI and type 2 diabetes, suggesting shared cardiometabolic contributions.

*Heart failure–varicose veins:*

Heart failure and varicose veins showed modest overlap (***r*_g_ = 0.21, SE = 0.07**), without additional supporting evidence beyond genome-wide correlation.

*Abdominal aortic aneurysm–aortic stenosis:*

AAA and AS showed no genome-wide genetic correlation but shared colocalising loci and FLAMES genes (*LPA/SLC22A3*), implicating lipoprotein-related pathways.

### Supplementary figures

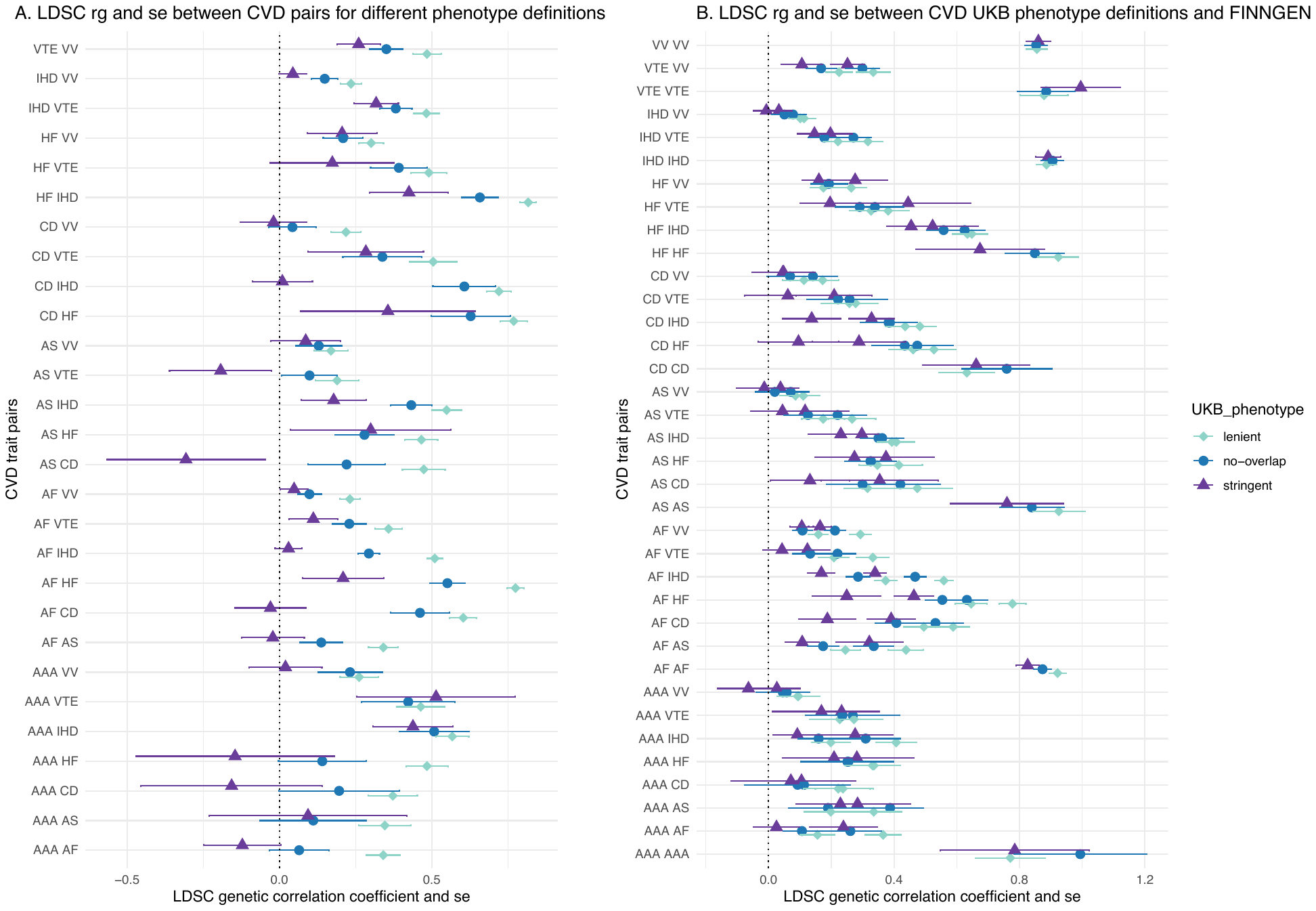

#### Supplementary Figure 1. Genetic correlations between CVD pairs under different case definitions.

*Forest plots show genetic correlations between all CVD pairs estimated using three alternative case definitions within the same biobank: lenient, stringent, and no–case-overlap (see Supplemental Note 2). Panel B shows the corresponding genetic correlations between each CVD definition and CVD GWAS from FinnGen. Points indicate point estimates and horizontal bars denote 95% confidence intervals. Two points are present per line in B due to two possible combinations of UKB and FinnGen phenotype for each pair (i.e., trait 1 (UKB) – trait 2 (FinnGen) and trait 1 (FinnGen) – trait 2 (UKB).*

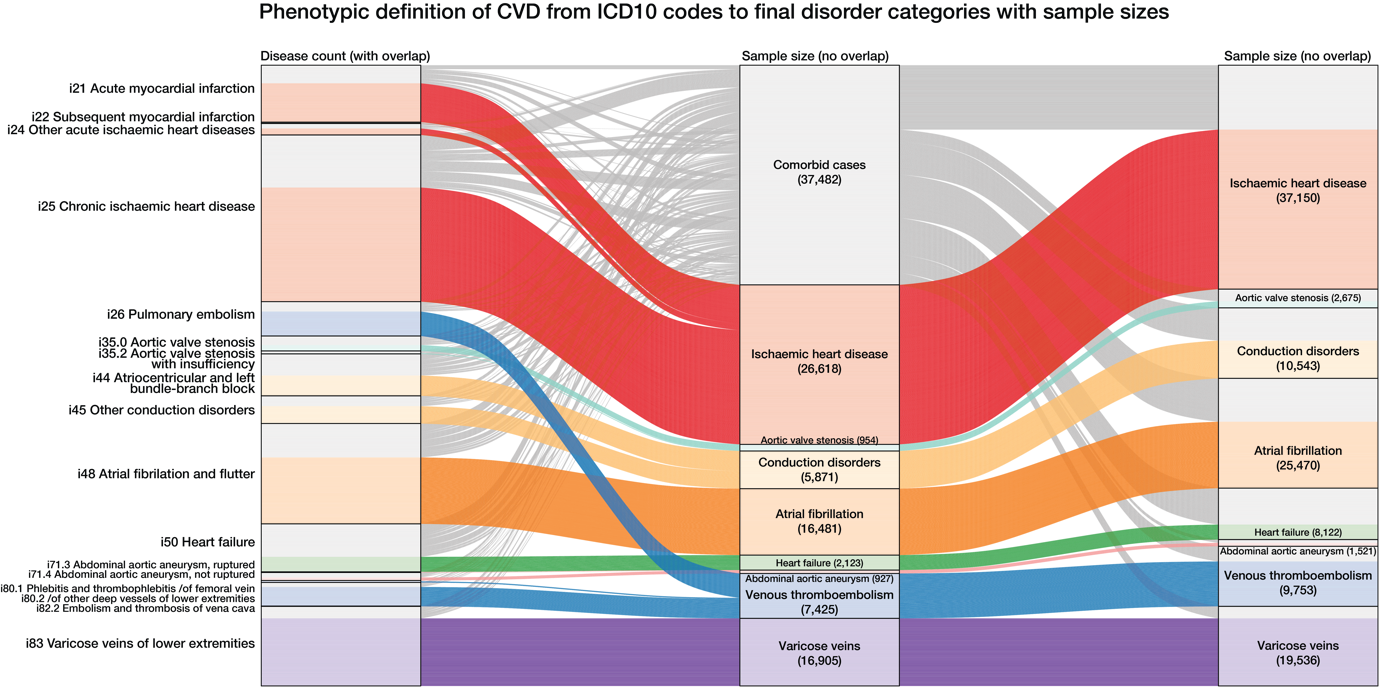

#### Supplementary Figure 2. Phenotype construction and handling of comorbid CVD cases.

*Alluvial diagram showing how ICD10 diagnosis codes are mapped to final CVD categories. The diagram illustrates identification of comorbid cases and their handling under the no–case-overlap approach, in which individuals with multiple CVD diagnoses are assigned to a single disease category and not included in analyses of their other diagnosed categories. Sample sizes are shown at each stage of phenotype construction.*

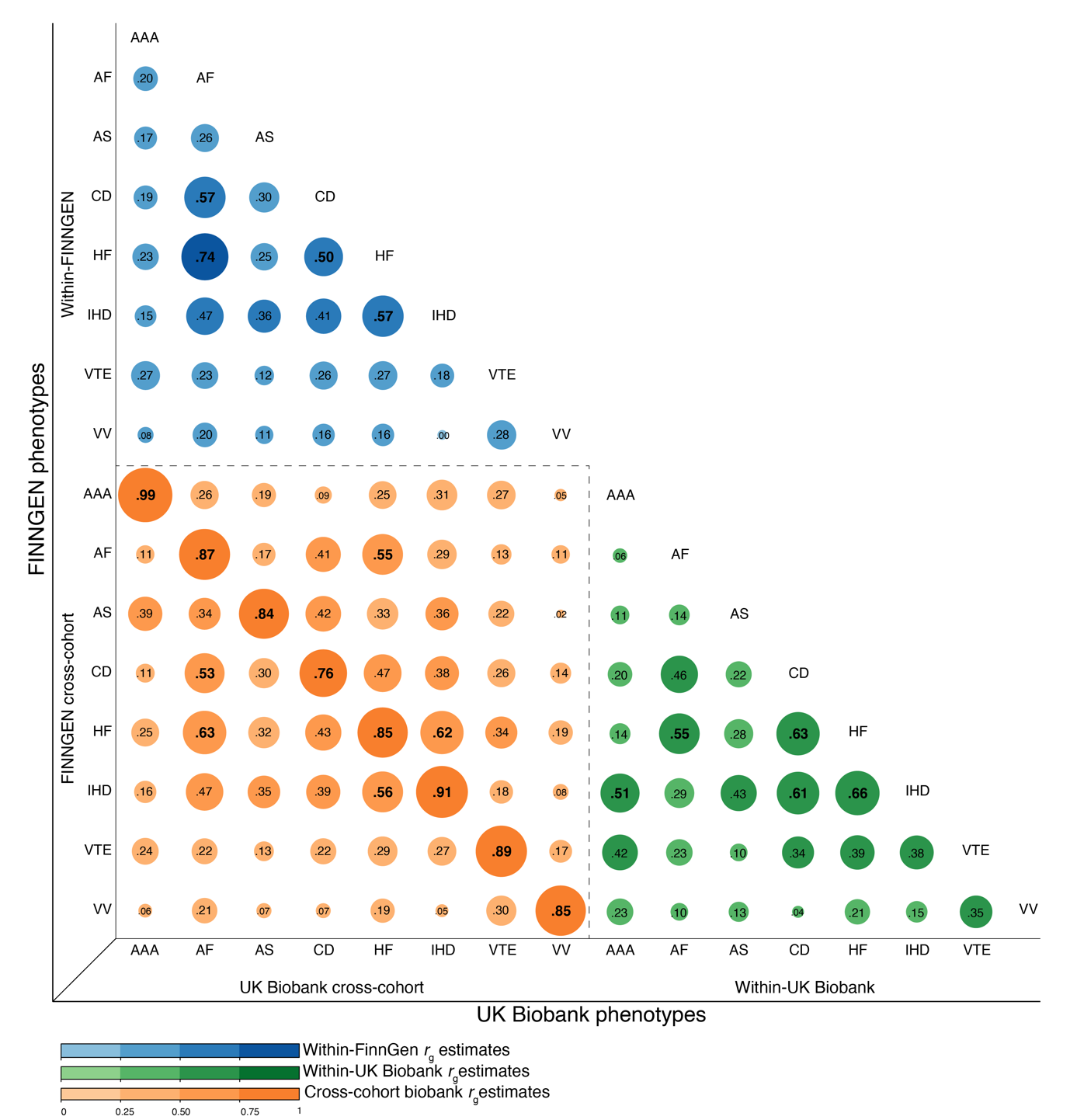

#### Supplementary Figure 3. Genetic correlations within and across biobanks (UKB and FinnGen)

*Genetic correlations within and between cohorts (UKB (no-case overlap definition) and FinnGen) showing LDSC estimates. Genetic correlation between-cohort of the same trait is generally high reflecting similarity in genetic liability captured of the GWAS.*

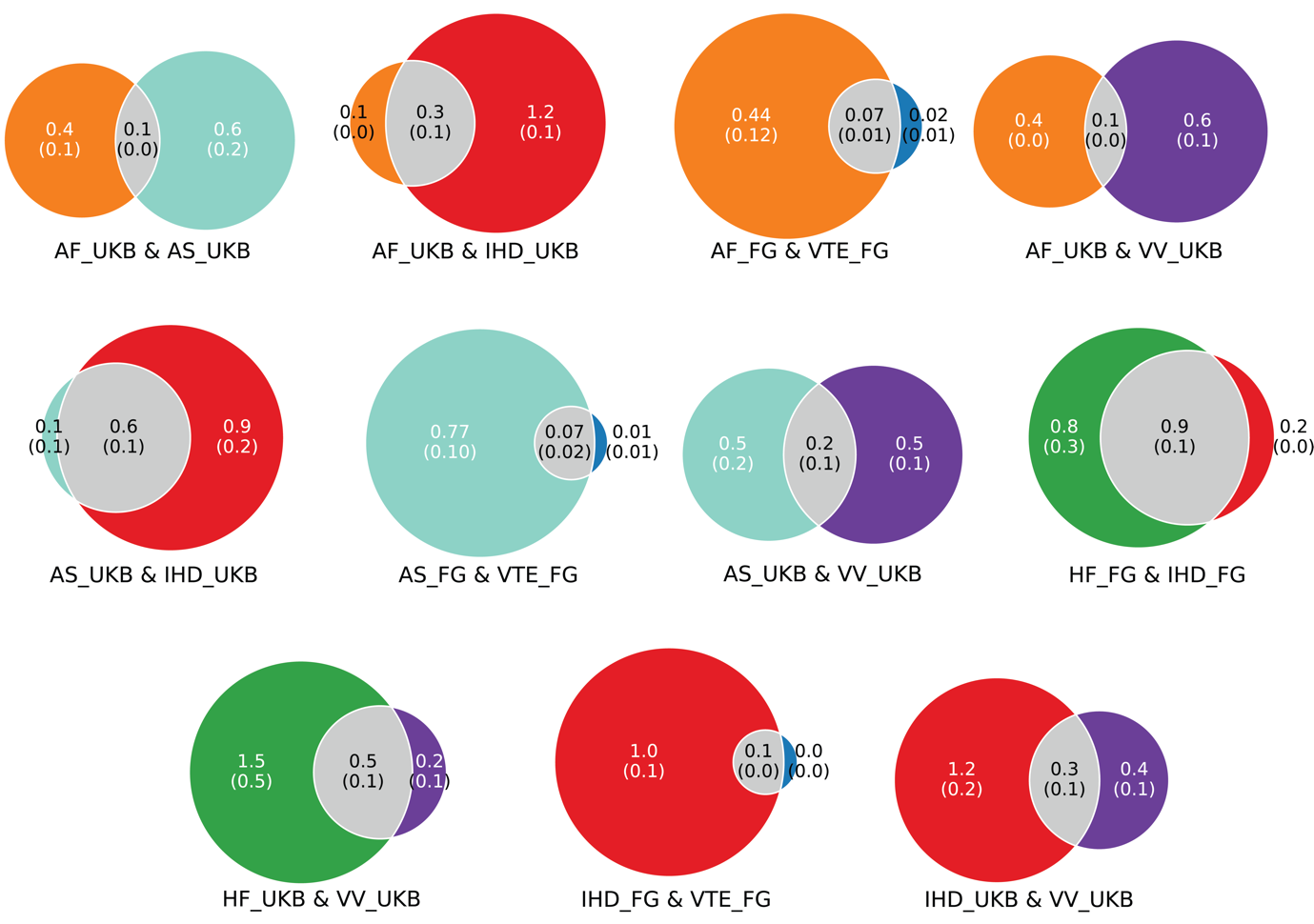

#### Supplementary Figure 4. Bivariate MiXeR showing Venn diagrams of CVD pair polygenicity

*Venn diagram of estimated bivariate MiXeR shared genetic architecture between CVD pairs.with sufficient model fit estimates (positive AIC). Traits with negative AIC are not reported above. Estimates are in 1000 SNPs, i.e., 0.6 (.1) denotes 600 shared SNPs with SE =100 between CVDs. MiXeR model fit was supported by AIC for all the presented CVD pairs. When UKB based CVDs did not achieve sufficient fit, but FinnGen CVD pairs did, the FinnGen CVD pairs were plotted instead.*

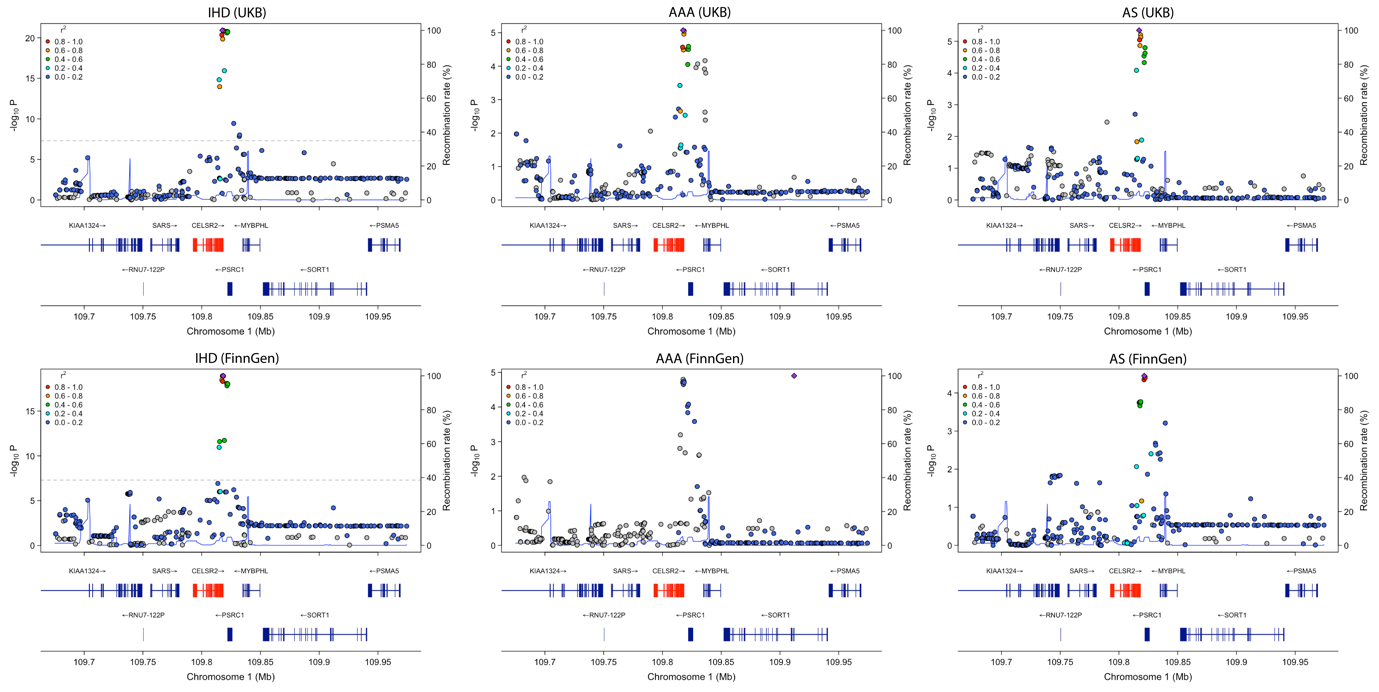

Supplementary Figure 5. Regional association plots for a shared locus on chromosome 1 across CVD traits**.** *Regional association plots showing genome-wide association signals at a shared locus on chromosome 1 (≈109.7–110.0 Mb) for IHD, AAA, and AS in UKB (top row) and FinnGen (bottom row). Points represent single-variant association statistics, colored by linkage disequilibrium with the lead variant. Gene models are shown below each panel, with the FLAMES-prioritized gene highlighted in red. LD estimates and locus construction were obtained using LDlink.*

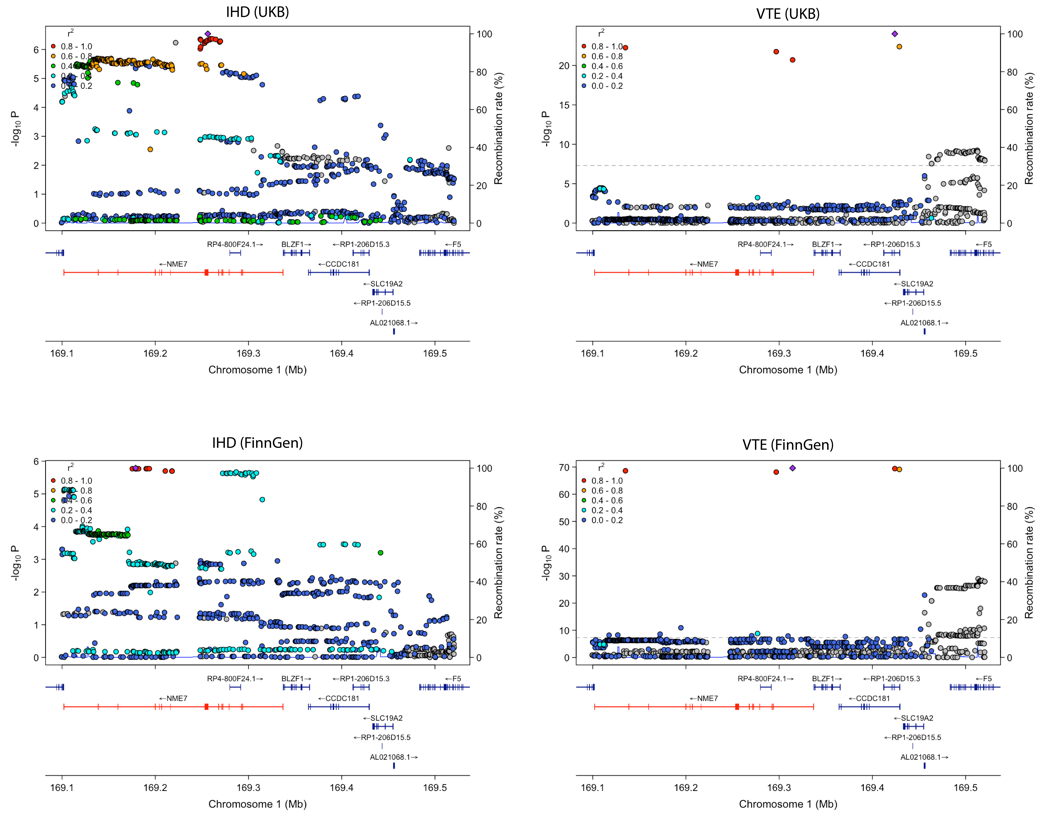

Supplementary Figure 6. Regional association plots for a shared locus on chromosome 1 across CVD traits**.** *Regional association plots showing genome-wide association signals at a shared locus on chromosome 1 (≈169.1–169.5 Mb) for IHD and VTE in UKB (top row) and FinnGen (bottom row). Points represent single-variant association statistics, colored by linkage disequilibrium with the lead variant. Gene models are shown below each panel, with the FLAMES-prioritized gene highlighted in red. LD estimates and locus construction were obtained using LDlink.*

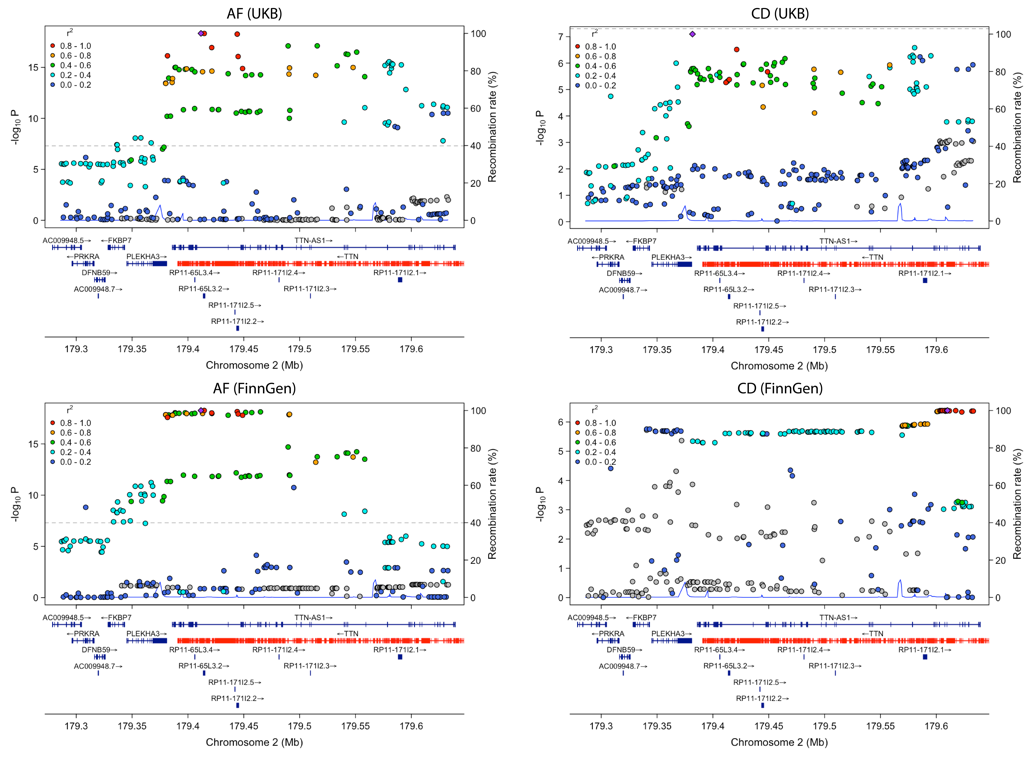

Supplementary Figure 7. Regional association plots for a shared locus on chromosome 2 across CVD traits**.** *Regional association plots showing genome-wide association signals at a shared locus on chromosome 2 (≈179.3–179.6 Mb) for AF and CD in UKB (top row) and FinnGen (bottom row). Points represent single-variant association statistics, colored by LD with the lead variant. Gene models are shown below each panel, with the FLAMES-prioritized gene highlighted in red. LD estimates and locus construction were obtained using LDlink.*

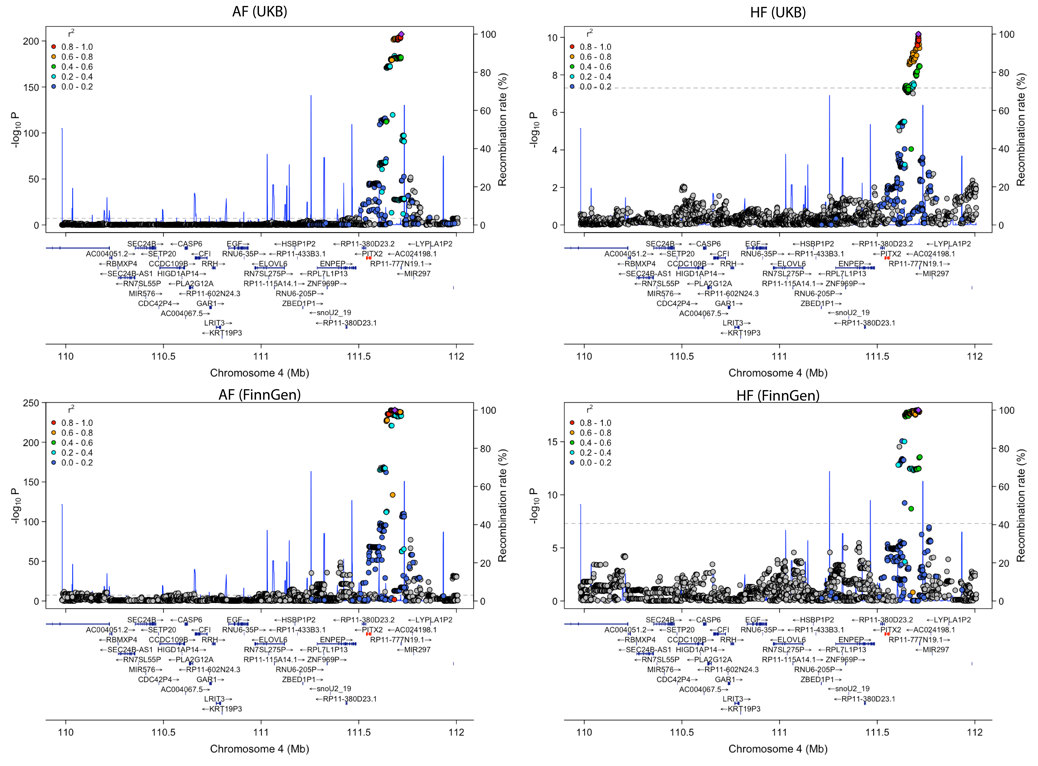

Supplementary Figure 8. Regional association plots for a shared locus on chromosome 4 across CVD traits**.** *Regional association plots showing genome-wide association signals at a shared locus on chromosome 4 (≈110.0–112.0 Mb) for AF and HF in UKB (top row) and FinnGen (bottom row). Points represent single-variant association statistics, colored by LD with the lead variant. Gene models are shown below each panel, with the FLAMES-prioritized gene highlighted in red. LD estimates and locus construction were obtained using LDlink.*

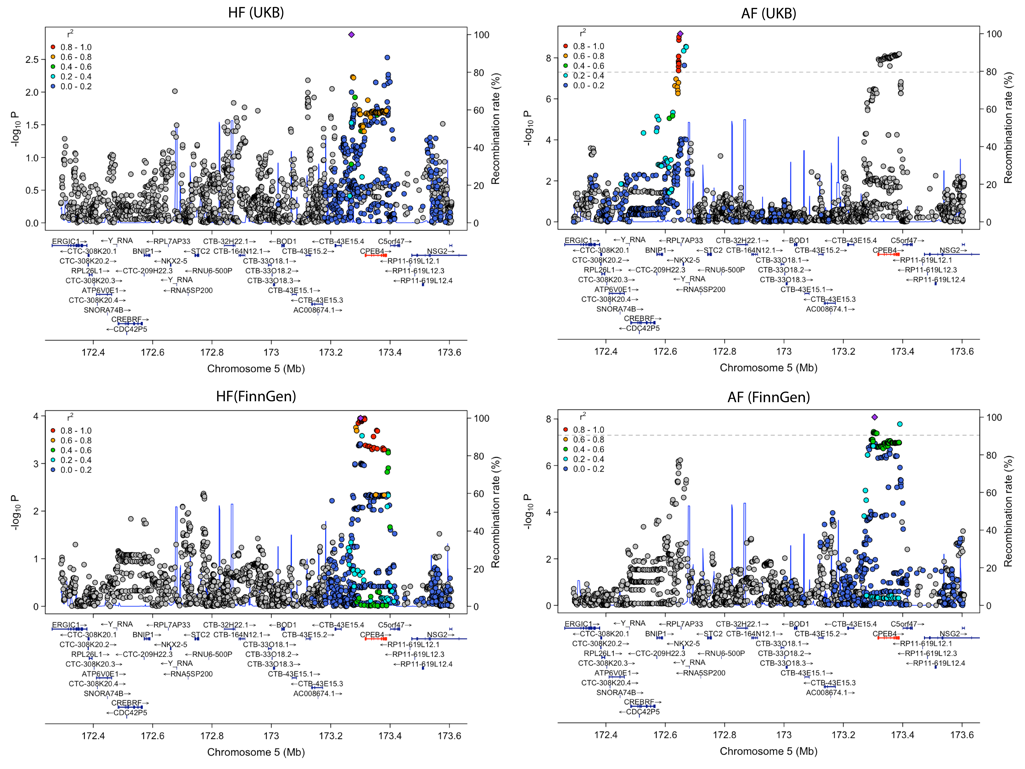

Supplementary Figure 9. Regional association plots for a shared locus on chromosome 5 across CVD traits**.** *Regional association plots showing genome-wide association signals at a shared locus on chromosome 5 (≈172.4–173.6 Mb) for HF and AF in UKB (top row) and FinnGen (bottom row). Points represent single-variant association statistics, colored by LD with the lead variant. Gene models are shown below each panel, with the FLAMES-prioritized gene highlighted in red. LD estimates and locus construction were obtained using LDlink.*

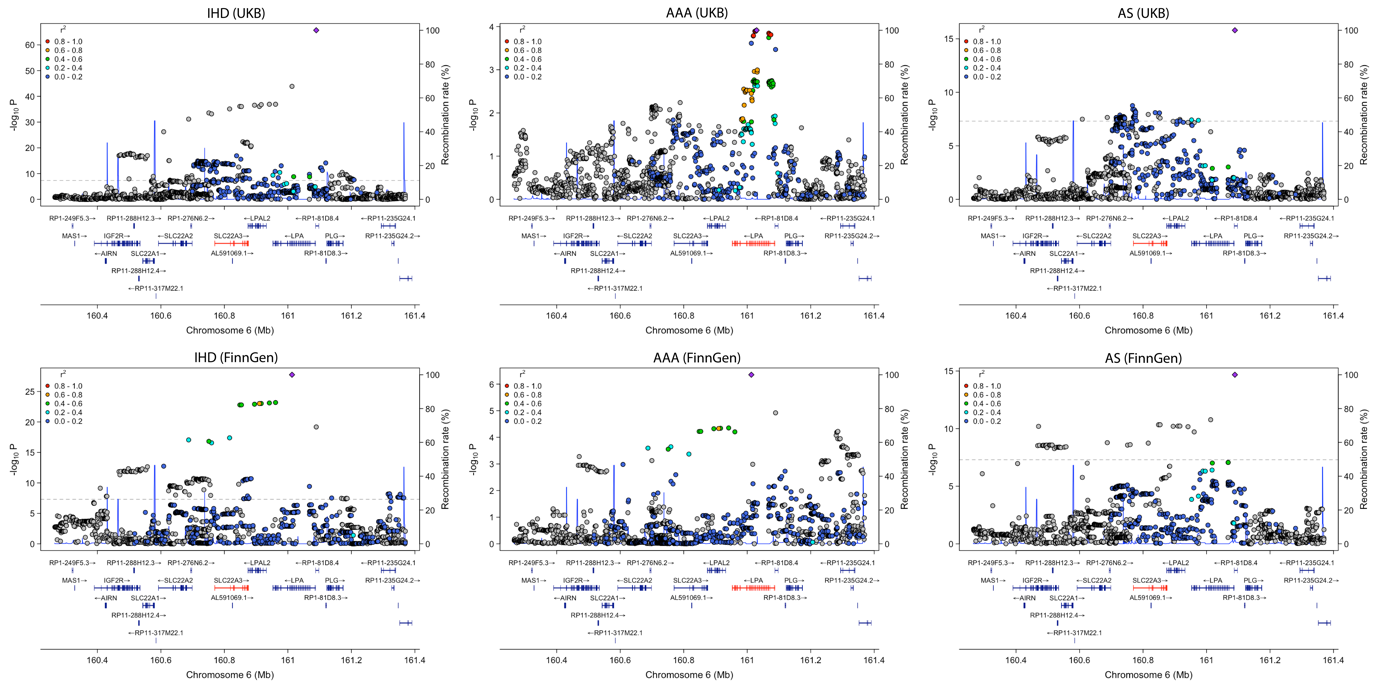

Supplementary Figure 10. Regional association plots for a shared locus on chromosome 6 across CVD traits**.** *Regional association plots showing genome-wide association signals at a shared locus on chromosome 6 (≈160.4–161.4 Mb) for IHD, AAA, and AS in UKB (top row) and FinnGen (bottom row). Points represent single-variant association statistics, colored by LD with the lead variant. Gene models are shown below each panel, with the FLAMES-prioritized gene highlighted in red. LD estimates and locus construction were obtained using LDlink*

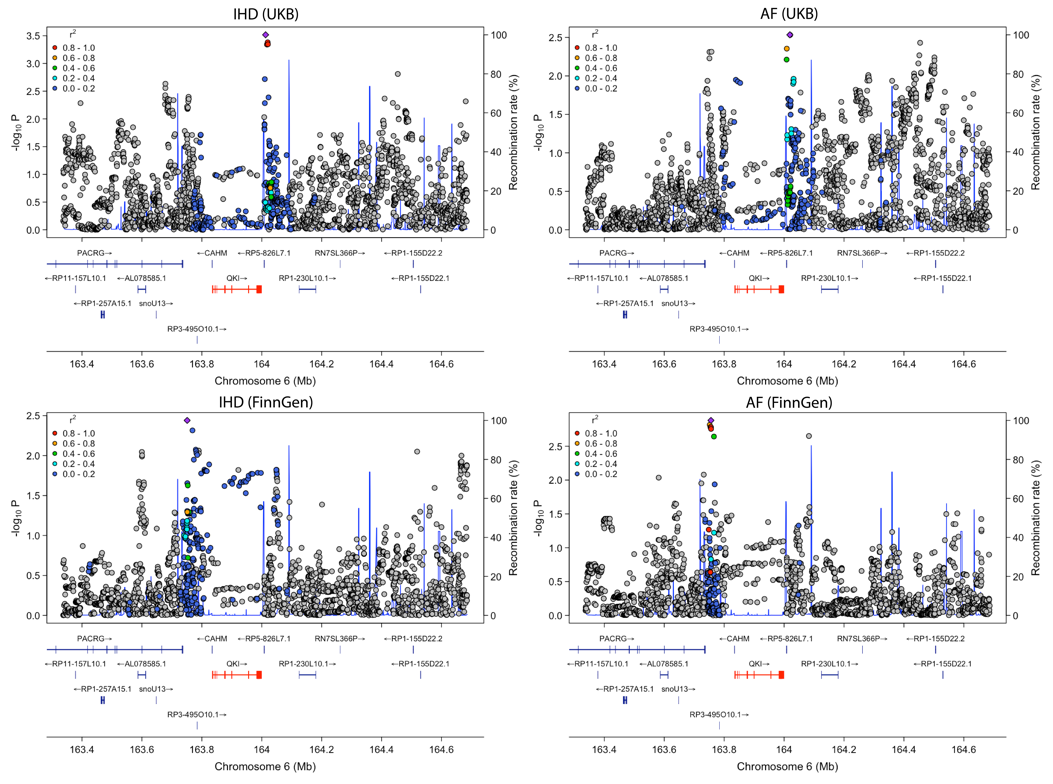

Supplementary Figure 11. Regional association plots for a shared locus on chromosome 6 across CVD traits**.** *Regional association plots showing genome-wide association signals at a shared locus on chromosome 6 (≈163.4–164.6 Mb) for IHD and AF in UKB (top row) and FinnGen (bottom row). Points represent single-variant association statistics, colored by LD with the lead variant. Gene models are shown below each panel, with the FLAMES-prioritized gene highlighted in red. LD estimates and locus construction were obtained using LDlink.*

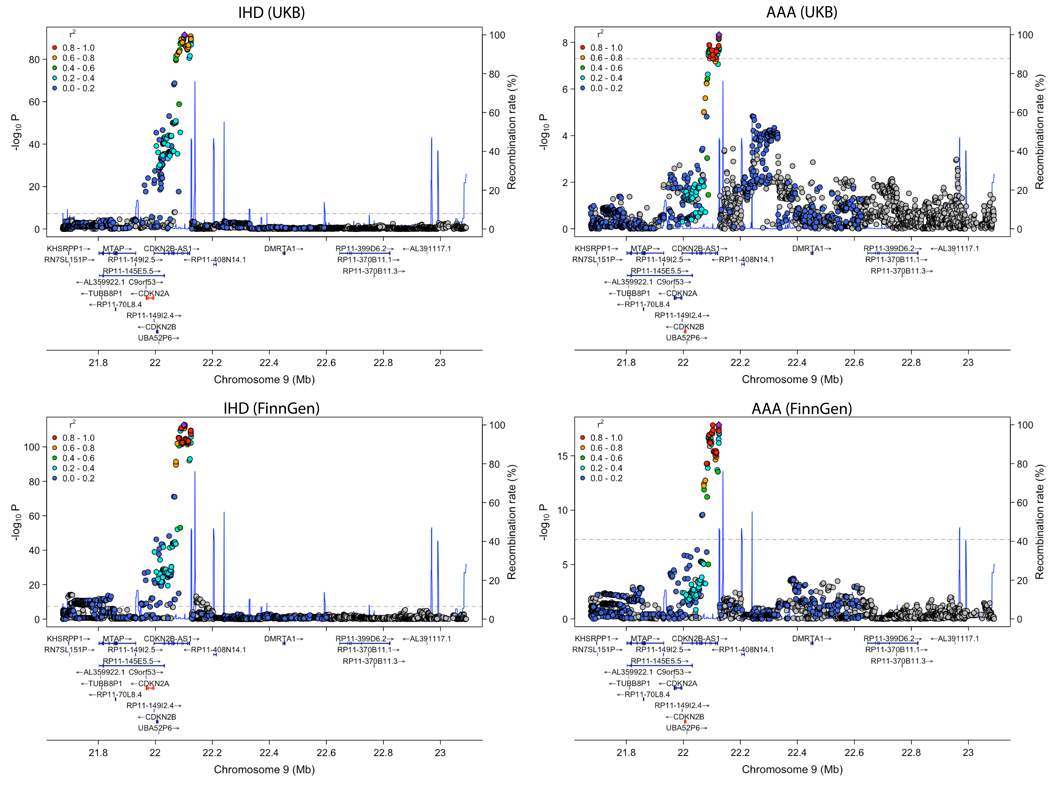

Supplementary Figure 12. Regional association plots for a shared locus on chromosome 9 across CVD traits**.** *Regional association plots showing genome-wide association signals at a shared locus on chromosome 9 (≈21.8–23.0 Mb) for IHD and AAA in UKB (top row) and FinnGen (bottom row). Points represent single-variant association statistics, colored by LD with the lead variant. Gene models are shown below each panel, with the FLAMES-prioritized gene highlighted in red. LD estimates and locus construction were obtained using LDlink.*

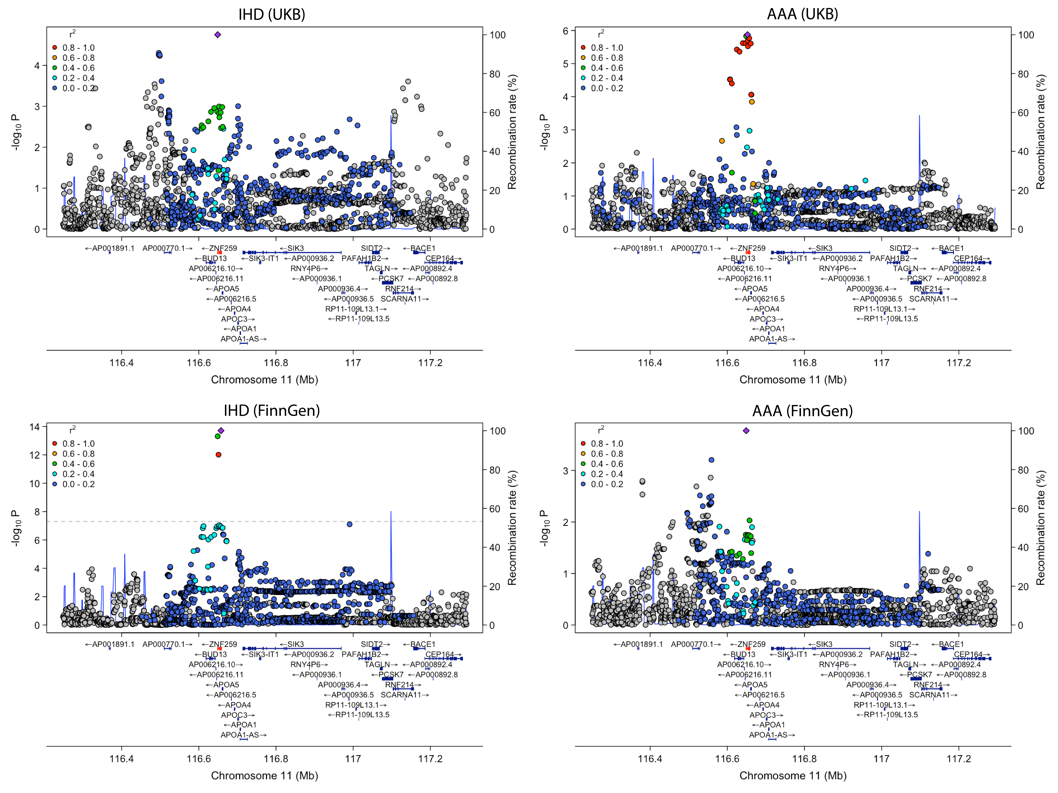

Supplementary Figure 13. Regional association plots for a shared locus on chromosome 11 across CVD traits**.** *Regional association plots showing genome-wide association signals at a shared locus on chromosome 11 (≈116.4–117.2 Mb) for IHD and AAA in UKB (top row) and FinnGen (bottom row). Points represent single-variant association statistics, colored by LD with the lead variant. Gene models are shown below each panel, with the FLAMES-prioritized gene highlighted in red. LD estimates and locus construction were obtained using LDlink.*

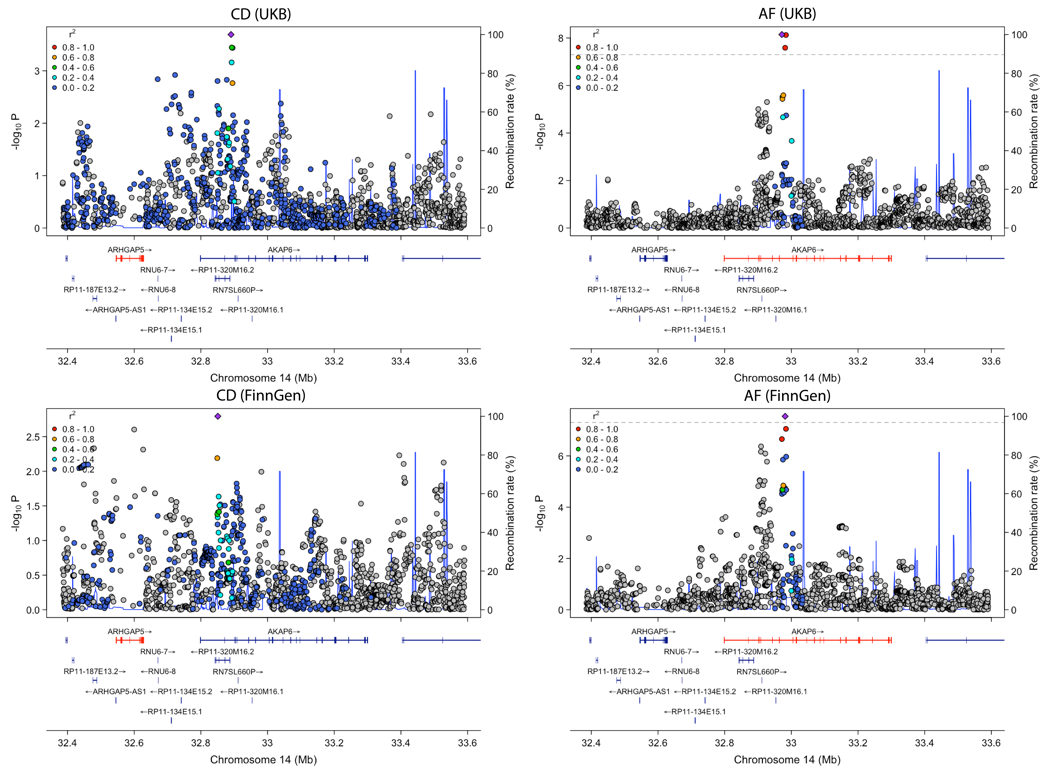

Supplementary Figure 14. Regional association plots for a shared locus on chromosome 14 across CVD traits**.** *Regional association plots showing genome-wide association signals at a shared locus on chromosome 14 (≈32.4–33.6 Mb) for CD and AF in UKB (top row) and FinnGen (bottom row). Points represent single-variant association statistics, colored by LD with the lead variant. Gene models are shown below each panel, with the FLAMES-prioritized gene highlighted in red. LD estimates and locus construction were obtained using LDlink.*

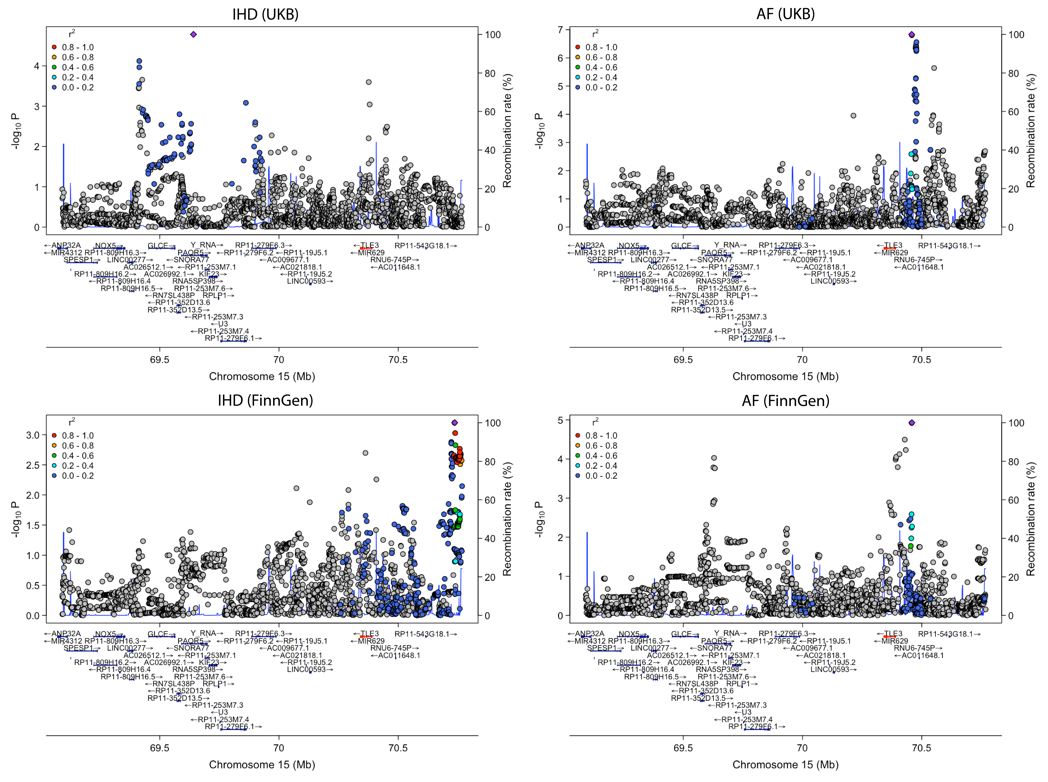

Supplementary Figure 15. Regional association plots for a shared locus on chromosome 15 across CVD traits**.** *Regional association plots showing genome-wide association signals at a shared locus on chromosome 15 (≈69.0–70.6 Mb) for IHD and AF in UKB (top row) and FinnGen (bottom row). Points represent single-variant association statistics, colored by LD with the lead variant. Gene models are shown below each panel, with the FLAMES-prioritized gene highlighted in red. LD estimates and locus construction were obtained using LDlink.*

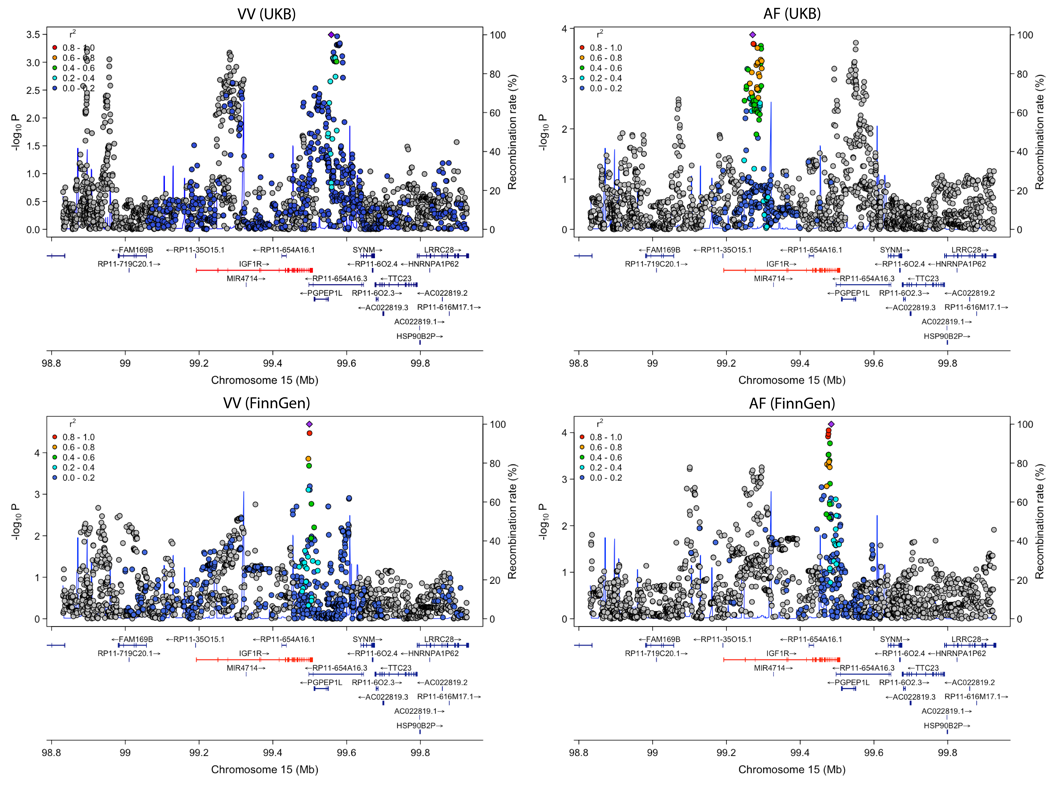

Supplementary Figure 16. Regional association plots for a shared locus on chromosome 15 across CVD traits**.** *Regional association plots showing genome-wide association signals at a shared locus on chromosome 15 (≈98.8–99.9 Mb) for VV and AF in UKB (top row) and FinnGen (bottom row). Points represent single-variant association statistics, colored by LD with the lead variant. Gene models are shown below each panel, with the FLAMES-prioritized gene highlighted in red. LD estimates and locus construction were obtained using LDlink.*

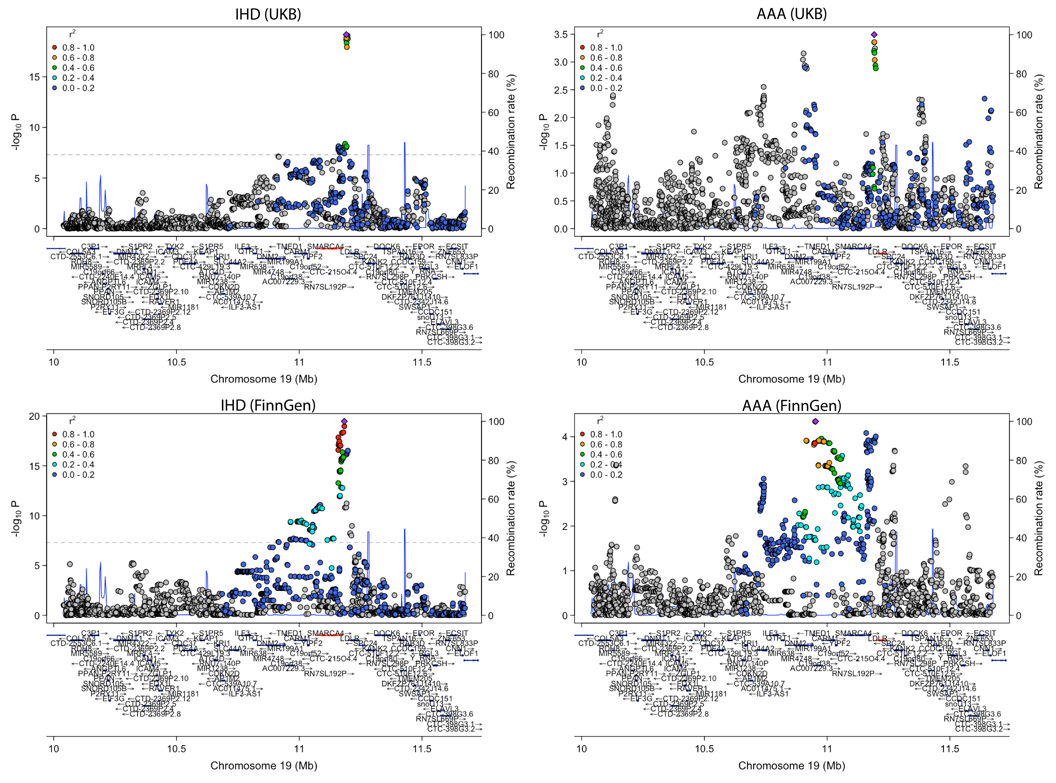

Supplementary Figure 17. Regional association plots for a shared locus on chromosome 19 across CVD traits**.** *Regional association plots showing genome-wide association signals at a shared locus on chromosome 19 (≈10.0–11.6 Mb) for IHD and AAA in UKB (top row) and FinnGen (bottom row). Points represent single-variant association statistics, colored by LD with the lead variant. Gene models are shown below each panel, with the FLAMES-prioritized gene highlighted in red. LD estimates and locus construction were obtained using LDlink.*

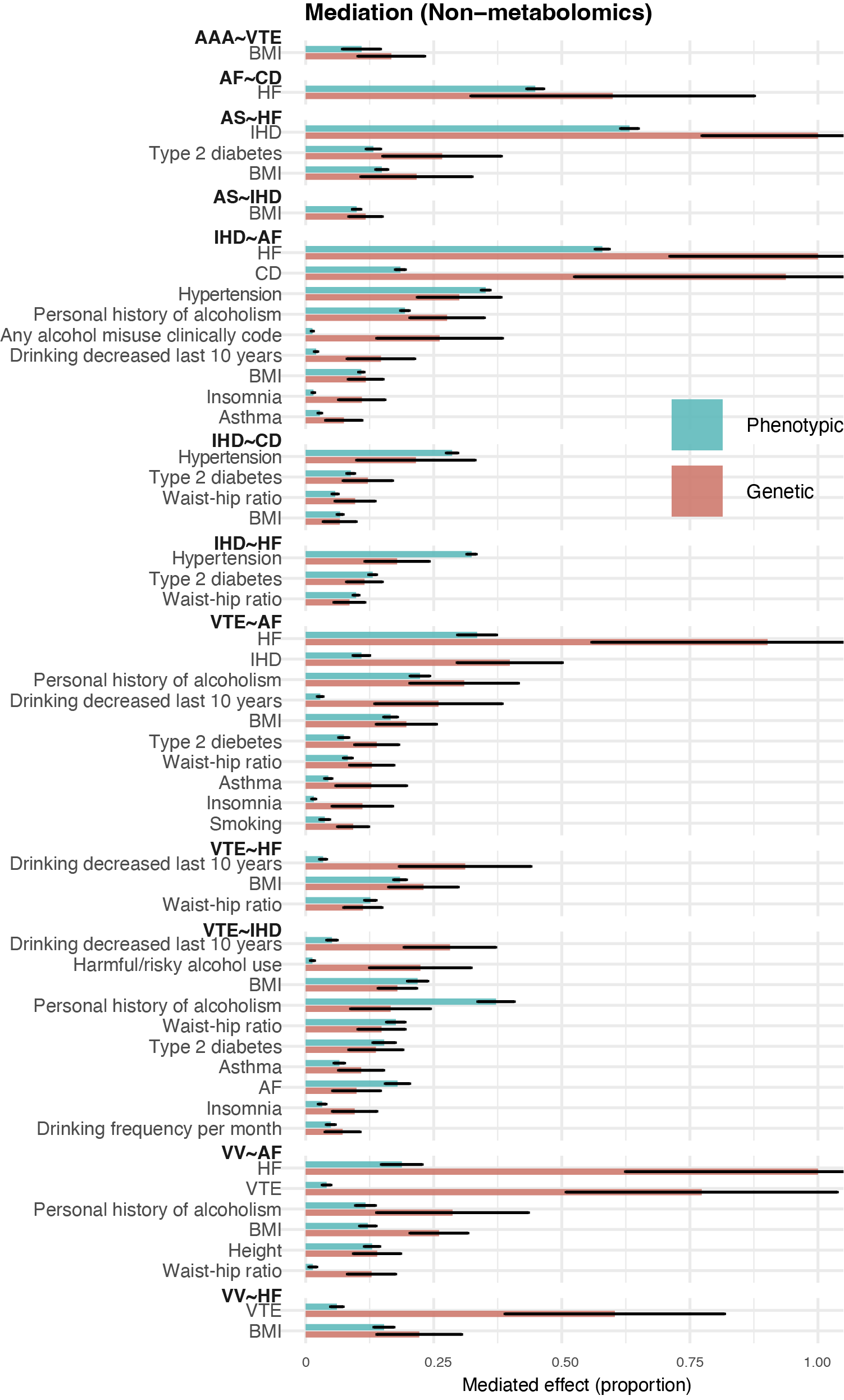

Supplementary Figure 18. Genetic and phenotypic mediation of CVD pair relationships by non-metabolic traits**.**

*Bar plots show the proportion of the association between CVD pairs mediated by non-metabolic traits, estimated at the phenotypic level (blue) and genetic level (red). For each mediator, mediation was estimated in both model orientations (A→B and B→A), and the minimum proportion mediated across orientations is shown as a conservative estimate. Error bars indicate standard errors.*

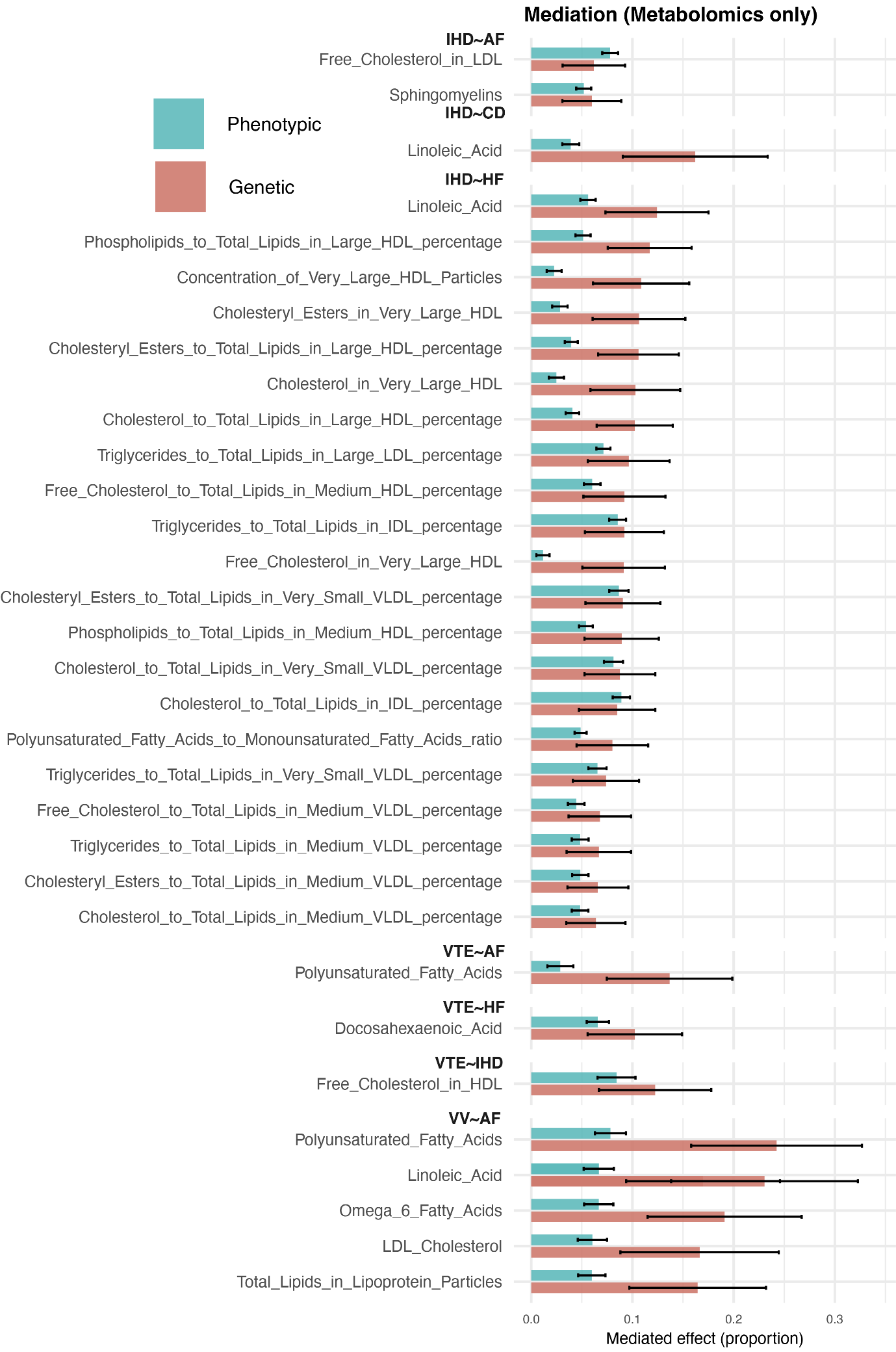

Supplementary Figure 19. Genetic and phenotypic mediation of CVD pair relationships by metabolomic traits. *Bar plots show the proportion of the association between CVD pairs mediated by circulating metabolomic traits, estimated at the phenotypic level (blue) and genetic level (red). For each mediator, mediation was estimated in both model orientations (A→B and B→A), and the minimum proportion mediated across orientations is shown as a conservative estimate. Error bars indicate standard errors.*

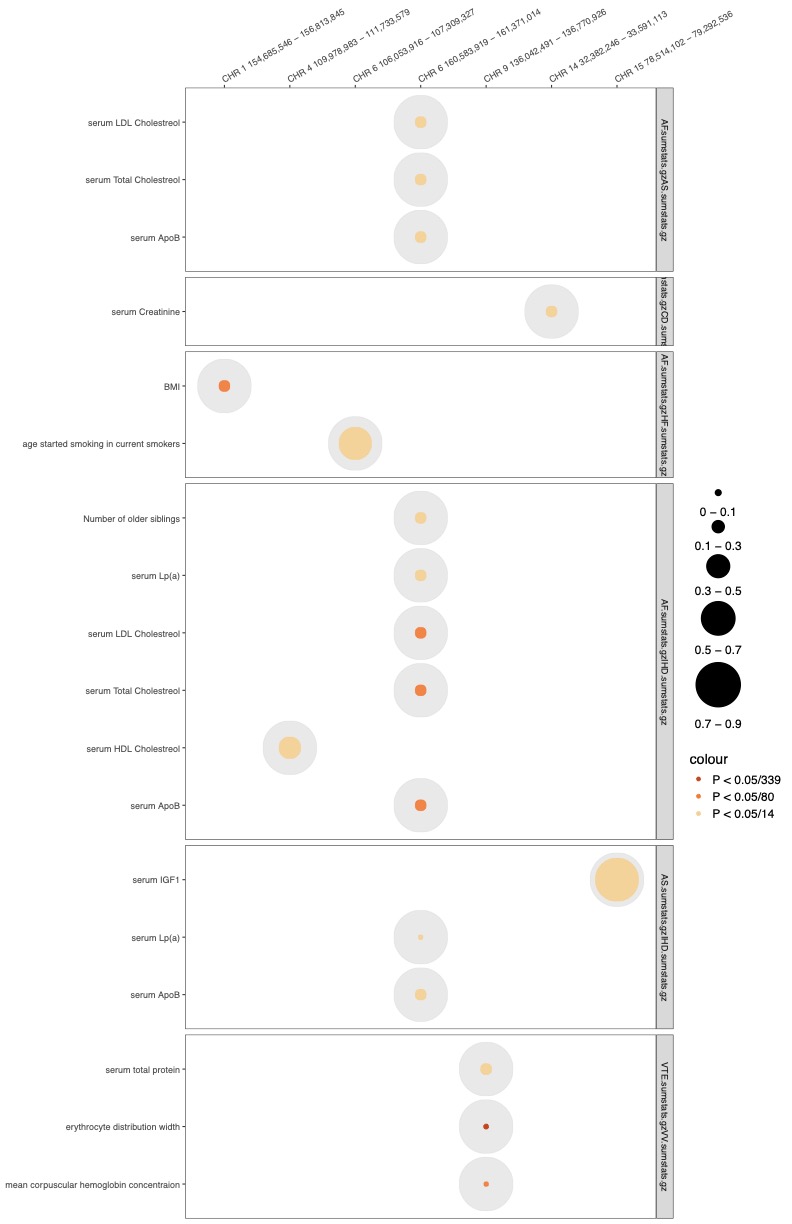

Supplementary Figure 20. Conditional local genetic correlation results for mediator traits across CVD pairs**.** *Bubble plot summarizing results from conditional local genetic correlation analyses for mediator traits showing nominally significant conditional effects. Rows indicate mediator traits, grouped by CVD pair, and columns indicate genomic regions. Circle size represents the estimated conditional effect, defined as the proportion of genetic covariance shared between the CVD pair that is also shared with the mediator trait. Gray circles indicate the theoretical maximum overlap, while colored circles show the observed overlap within each genomic region. Color denotes statistical significance of the conditional effect based on increasing levels of multiple-testing correction, as indicated in the legend. Genomic regions are labeled at the top of the plot.*
